# Digital Screener of Socio-Motor Agency Balancing Autonomy and Control

**DOI:** 10.1101/2023.10.25.23297428

**Authors:** Theodoros Bermperidis, Richa Rai, Elizabeth B Torres

## Abstract

Dyadic social interactions evoke complex dynamics between two agents that while exchanging unequal levels of body autonomy and motor control, may find a fine balance to take turns and gradually build social rapport. To study the evolution of such complex interactions, we currently rely exclusively on subjective pencil and paper means. Here we complement this approach with objective biometrics of socio-motor behaviors conducive of socio-motor agency. Using a common clinical test as the backdrop of our study to probe social interactions between a child and a clinician, we demonstrate new ways to streamline the detection of social readiness potential in both typically developing and autistic children. We highlight differences between males and females and uncover a new data type amenable to generalize our results to any social settings. The new methods convert dyadic bodily biorhythmic activity into spike trains and demonstrates that in the context of dyadic behavioral analyses, they are well characterized by a continuous gamma process independent from corresponding binary spike rates. We offer a new framework that combines stochastic analyses, nonlinear dynamics, and information theory, to facilitate scaling the screening and tracking of social interactions with applications to autism.

## 1 Introduction

All research involving autism is (arguably) fundamentally tied to the Autism Diagnosis Observation Schedule (ADOS, currently in version 2 [1; 2; 3; 4].) Research spanning disparate fields, from genomics to complex social interactions relies on this test as the gold standard to classify humans across the lifespan as autistic or autism spectrum. Although clinically validated, the ADOS-based diagnosis misses females [5; 6; 7]. Moreover, there are not enough raters to absorb the large number of toddlers, children, and adults that according to various screening tools, are suspected as autistic today. The test is long and taxing on both the child and the clinician administering it because it has an average of 27 tasks aimed at engaging the child through social presses and expecting overtures from the child.

The ADOS is a dynamic and flexible test in the sense that the clinician can choose the tasks according to the flow of the child’s performance. It also adapts the test on demand, choosing the module that best agrees with the child’s communication level. However, the interaction occurs while the clinician also scores the child’s performance. Though valid to probe social competence, many of the tasks artificially rob the child of a chance to be naturally social, as the interaction is also taxing on the clinician and at times, awkward and seemingly forced. In this sense, several of the tasks might be biased, interfering with the child’s agency, and robbing the clinician of the spontaneity characteristic of a natural social exchange. In this sense, we need objective ways to quantify this potential bias that such a taxing effect may produce on both social agents.

Prior work analyzing thousands of ADOS score records found non-obvious issues with the statistical foundations used to validate this test. While there are theoretical requirements of normality and homogeneous variance in the signal detection theory used to validate the ADOS [8; 9], as these assumptions are required for independence between bias and sensitivity [10], the empirical data across thousands of records, violate these assumptions [10]. New methods have then been proposed to help reduce the number of tasks [11], while also utilizing motor signatures to identify females [4; 11; 12; 13]. However, there are no means to define naturalistic social agency in the dyad and to identify tasks that enhance it. Furthermore, no means to implement these tasks using artificial intelligence (AI) and machine learning (ML) methods have been proposed. Such approaches would help us speed up, automate, and scale the assessment process, particularly doing so with respect to currently underdiagnosed females [5].

We reasoned in the present work that the digital ADOS [11], *i.e*., the ADOS that is digitally recorded while the child and clinician interact, could leverage the validity of this test as the gold standard for clinical and research use, while providing a streamlined version of it that could help us (1) identify objective biometrics of social agency and (2) automate the process of identifying socially compliant tasks using methods from Artificial Intelligence (AI) and Machine Learning (ML).

In our approach, socially compliant tasks are those which provide social agency to the child that is being diagnosed. More precisely, we here define social agency as the balance between autonomy and control during a social exchange. Autonomy is defined as the ability of the child to lead the conversation as much as the clinician does, rather than always following the lead of the clinician. Control is defined as the ability of the child to effectively predict the consequences of impending social actions and overall behaviors, based on intact motor control. Both concepts are illustrated in Figure 1.

**Figure 1.**
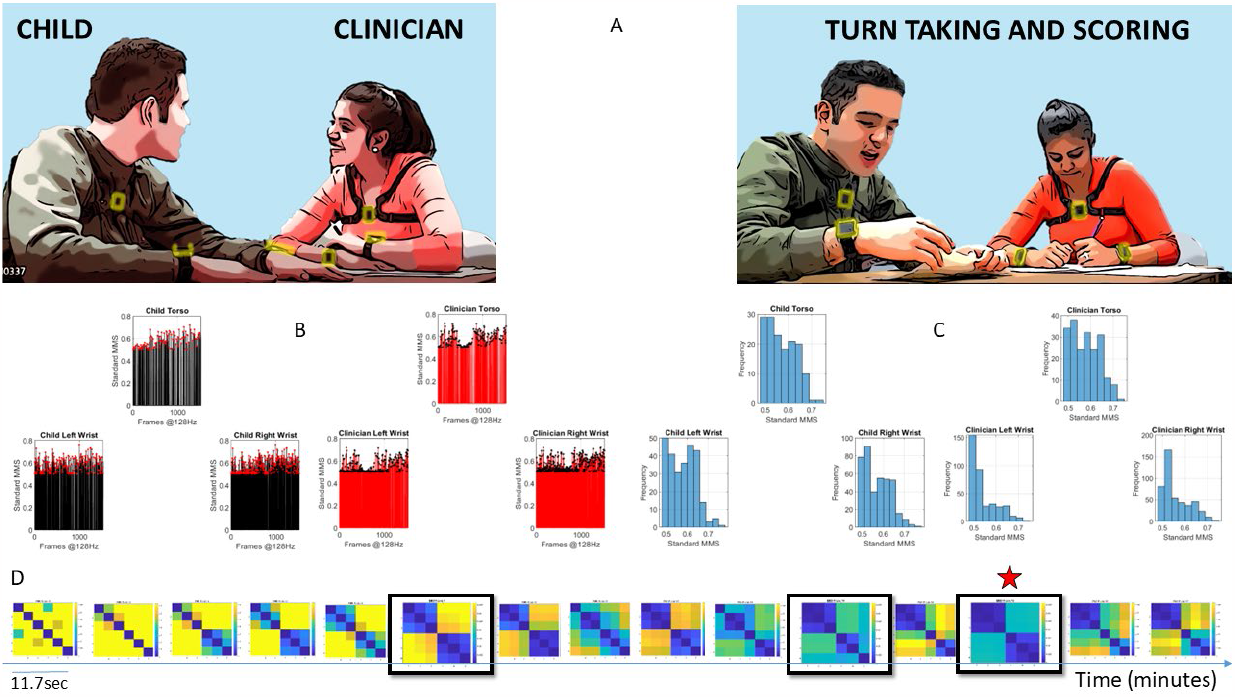
Digitally characterizing rapport and turn taking during an ADOS-based social interaction. (A) Snapshots of the interaction between a child and clinician wearing 6 biosensors, 3 on each body, synchronously registering motion at 128Hz. Clinician-led interaction and note taking while rating the interaction. (B) Sample standardized micromovement spikes (MMS) derived from angular speed capturing approximately 11.7seconds of social exchange. (C) Frequency histograms of the MMS peaks, (one frame) from each sensor on the child and clinician. (D) Pairwise comparison of the histograms evolution using the Earth Movers’ distance similarity metric. Entries reflect the 6×6 matrix (child and clinician, 3 sensors at the torso, right and left wrist) as in (A). Off diagonal entries are the shared dyadic space, while entries next to the diagonal are the child’s or clinician’s activities in standalone mode. Blue to yellow color EMD scale ranks from most to least similar spike patterns. Star marks the maximal similarity.

Socio-motor agency can be impeded if neurodevelopment undergoes a different maturation path [3; 14; 15]. If the child, for example, has excessive motor noise and motor randomness in its performance, the predictive ability required for self-motor control will be compromised [3] and with it, the overall control ability will be altered. This alteration will also in turn affect the clinician’s perception of the child’s nuanced micro-motions underlying social behaviors, thus biasing the assessment [3; 11; 14]. Under such circumstances, socio-motor agency can be impeded, as can be the rating of the child by the clinician. Therefore, the tasks that manifest excess random noise of the joint dyadic motor patterns (lower control of the dyad) and / or excess lead of the clinician within the dyad (lower autonomy of the child), are inevitably bound to bias the clinician’s scoring towards a deficit model of autism. In contrast, the tasks that manifest high dyadic control and autonomy of the child are bound to boast social agency, according to our biometric definition. These tasks can provide a more appropriate model of readiness potential for social exchange. While detecting a problem relative to normative data, this new model can also do so in a fair, unbiased manner. In this sense, the child has a chance to succeed. In turn, the clinician can presume competence and identify areas of strength to recommend treatments more appropriately. Such treatments will rather be grounded on the non-obvious, nuanced aspects of behaviors occurring at a micro-level that escapes the naked eye. Yet they will be quantifiable with biosensors that read out biorhythmic activities from the nervous systems with sub-second resolution.

We here introduce a theoretical framework grounded on empirically derived power (scaling) laws of human ontogenetically orderly (neurodevelopmental) maturation. This framework connects stochastic analysis of human biorhythmic (time series) data with information theoretical metrics. We define new truly personalized computational indexes of dyadic control, autonomy, and socio-motor agency from biosensors’ digital data using as guidance the digitized ADOS-2. Then, we identify socially compliant tasks *i.e*., ADOS-2 tasks with balanced socio-motor agency, thus streamlining the digital ADOS-2. Lastly, we propose new ways to help automate and speed up autism screening and detection based on these socially appropriate tasks identified from the motor variability of the interactive dyad, rather than from the child’s or the clinician’s performance alone.

## 2 Methods and Analyses

### 2.1 Participants

A total of 29 children including 19 males and 10 females spanning 4-15 years of age and two adult clinicians participated in the study (See Table 1). Children participated in multiple sessions over the span of 2 ½ years with one clinician per session and were administered a specific module per session, *i.e*., a specific subset of ADOS tasks, in accordance with their age, level of development and spoken language.

**Table 1.**
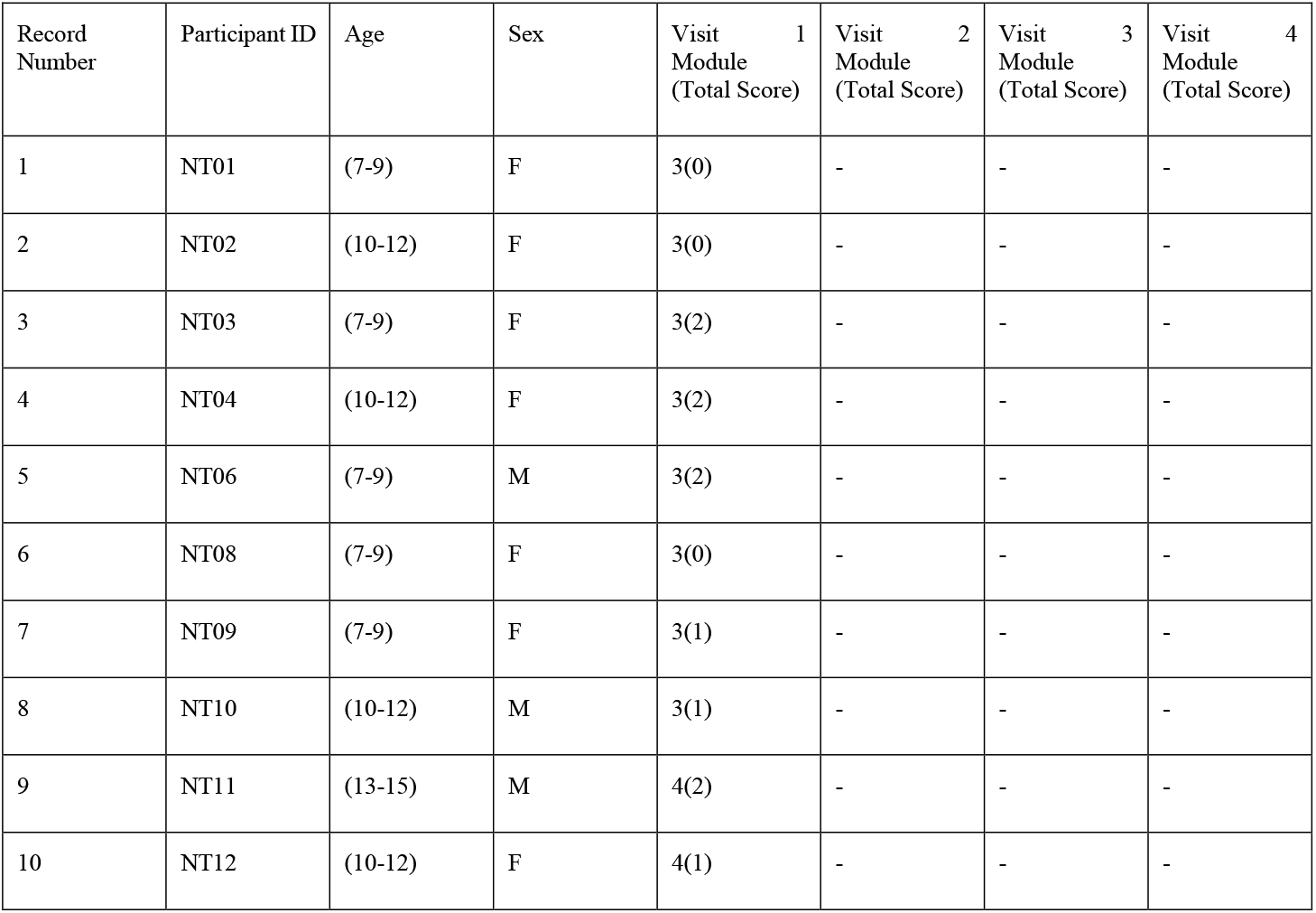
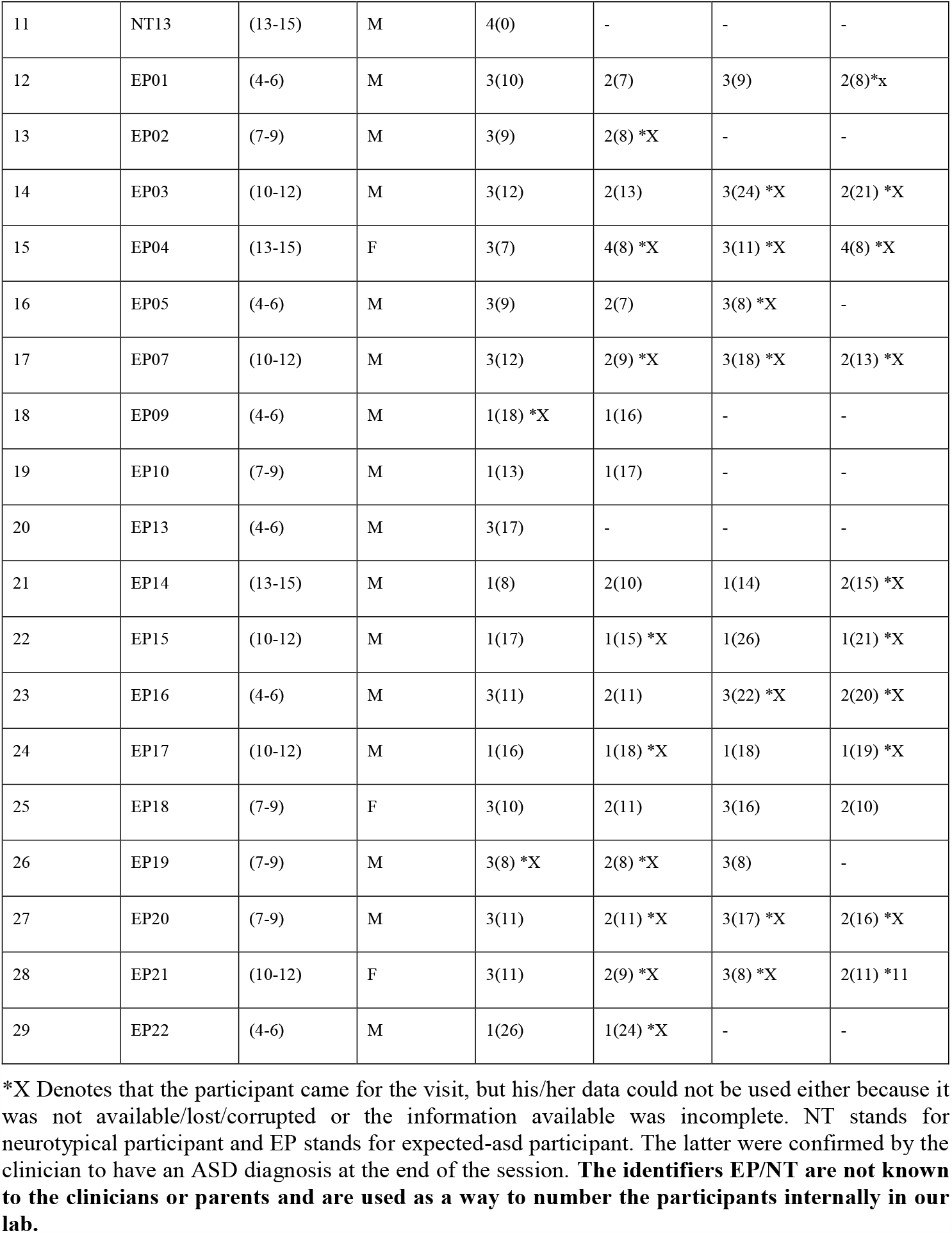
Participants information.

### 2.2 Raw Data Acquisition

Digital data were acquired during each session using light wearable sensors (APDM Opals, Portland, OR, USA). Six sensors were used, two on the left and right wrist and one on the torso, both on the child and clinician. The sensors continuously and synchronously recorded triaxial accelerometry and gyroscopic data at a sampling frequency of 128 Hz. The recording environment followed the standardized ADOS requirements using similar table and sitting arrangements for the clinician-child dyad. The two clinicians were unaware of the goals of the study.

### 2.3 Data Type: The Micro-Movement Spikes Derivation

Scalar values of angular speed from orientation data that the gyroscopes recorded (or acceleration from the inertial measurement units) were acquired using the Euclidean norm (using Equation 2.1) of the coordinate components of motion as measured by the sensors:

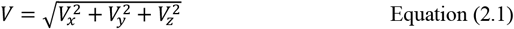

From here onward, all analyses refer to the scalar value V, the angular speed in deg/sec.

Human motion data such as angular speed and acceleration, are inherently biased by allometric effects and anatomical differences across subjects. To scale out such artifacts, we normalize motion data fluctuations (peaks and valleys) (using Equation 2.2) as relative deviations from the empirically estimated Gamma mean. The mean is estimated by fitting the continuous Gamma family of distributions to the raw peak data. The Gamma family of distributions has been consistently found to be the best candidate to fit human peak activity motion data, according to Maximum Likelihood Estimation (MLE). After we shift and center our data around the Gamma mean, we scale and map peak motion activity to the [0,1] interval according to the local minima average:

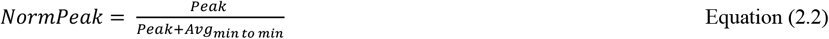

The normalized peak series, called Micro Movements Spikes (MMS)™ conserve the temporal structure of the original speed/acceleration time series. They represent “*quiet*” times interspersed with bouts of activity away from mean activity.

### 2.4 The Gamma Process of the MMS

The normalized speed MMS are best fit (in the MLE sense) by the continuous Gamma family of probability distributions [3; 16]. Furthermore, the parameters of the Gamma distribution, shape k and scale θ have been found across multiple studies from our laboratory, including the present one (see Results), to follow a Power Law of the form described in Equation 2.3:

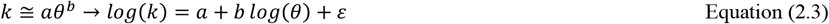

Where ε is a small error term and b < 0. This Power Law for the standardized MMS time series reveals a maturation process of the motor code for voluntary [3; 17] and involuntary [18] motions. This law is very important because it provides us with a quantitative framework to interpret fluctuations in biorhythmic data that range from random to predictive.

Importantly, the continuous Gamma family of probability distributions has the first (mean) and second (variance) moments expressed in terms of the shape and scale described by Equations 2.4)

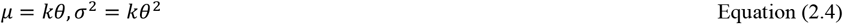

Then, using Equations 2.4, the Noise-to-Signal Ratio (NSR) of the MMS reduces to the Gamma scale parameter as in Equation 2.5:

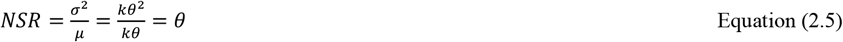

The Gamma scale parameter in Equation 2.5 fully characterizes the noise of the motor patterns of the interactive dyad (or of the participant), *i.e*., in relation to the level of fluctuations of angular speed during the ADOS activities.

Empirical estimation of these parameters in thousands of participants over a decade of work with humans along the lifespan, and across disorders of the nervous system, has revealed an interpretation for the Gamma log-log parameter plane. Distributions that fall along high NSR regimes are also close to the memoryless random regime of the special exponential distribution case (when k = 1). Points in mid NSR correspond to heavy-tailed Gamma distributions. Then, low NSR (or high signal = 1/NSR) are congruent with symmetric shapes (Gaussian-like) distributions. High-signal Gaussian regimens are highly predictable in contrast to High-noise memoryless random Exponential regimes. As such, this parameter plane is empirically interpretable.

### 2.5 Quantifying Motor Control from the Perspective of an Agent

Noise-to-Signal Ratio measures the degree of motion variability away from mean activity. Small NSR characterizes steady and smooth motion, akin of goal-oriented behavior, as experienced from the perspective of the agent/ child. On the other hand, a high NSR indicates unpredictable and random motion. In that sense, the NSR is a proxy for motor control and quantifies the existence of predictable motor patterns. Because the NSR is calculated on the standardized MMS, motor noise does not depend on the anatomy of the individual as it is scaled by the mean amplitude of motion.

### 2.6 An Information Theoretic Approach to the Analyses of the MMS

The presence of MMS peaks indicates an outburst of activity away from baseline. This is informative of a motor activity. When we also consider the temporal distribution of MMS peaks, a train of such spikes can be viewed as a representation of information regarding human motion variability through time. When we consider multiple sensors sampling in synchrony, the MMS spikes carry spatiotemporal information about the bursts of distributed bodily activity in the motor system.

### 2.7 Binary Trains of MMS

If we transform the MMS data so that the presence of a peak corresponds to the binary “1” and an absence of a peak corresponds to the binary “0”, we can represent normalized speed (or acceleration) as a stochastic binary sequence. An underlying mechanism stochastically generates bursts of activity, and this is equivalent to randomly generating 0s and 1s from an underlying binary alphabet as in Equation 2.6.

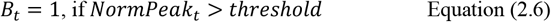

Let’s assume that *B*_*t*_ is a random sample drawn from an underlying probability distribution at time *t*.

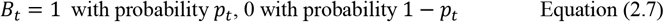

Entropy H in Equation 2.8 is an information theoretic measure that quantifies the amount of information in a random variable that follows a probability distribution *P*_*x*_ [19] and is equal to:

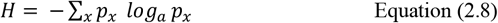

In the case of the binary process, the amount of information of the random variable of an activity outburst (MMS) is given in Equation 2.9:

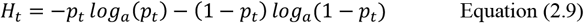

Which takes the maximum value of 1 when *p*_*t*_ *=* 0.5 and the minimum value of 0, when *p*_*t*_ *=* 0 or 1. Intuitively, entropy measures either the uncertainty regarding the outcome of a random realization of the random variable before that variable is measured or equivalently, the amount of information we get when we observe the variable. If we know for examples, that with a 100 % chance *B*_*t*_ *=* 1, the entropy is zero as we have no uncertainty about the outcome of the measurement, and no valuable information is provided to us. However, if with a 50 % chance *B*_*t*_ *=* 1, the entropy is at its maximum because we are totally uncertain whether the outcome will be 0 or 1 and observing the outcome gives us maximal information, specifically, 1 bit of information in the case of a base 2 logarithm (*N =* 2).

### 2.8 Measuring Randomness *vs*. Predictability Using Entropy Rate

The definition of entropy can be generalized for the case of multiple random variables *X*_1_, *X*_2_, …, *X*_*N*_, as in equation 2.10, by considering the joint probability distribution 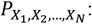:

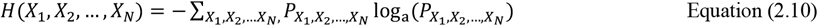

In the case of a stationary stochastic process *X* (*i.e*., statistical properties preserved over time) which takes values from a discrete alphabet *K* (in the case of the binary MMS), we can define the entropy rate of the process as in Equation 2.11:

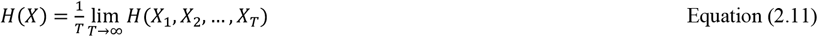

This quantity measures how much the process changes over time, *i.e*., the information that is carried in a new value. It measures the degree of randomness (unpredictability) of the underlying dynamical system [19; 20; 21; 22].

### 2.9 Randomness for Dynamical Systems

The concept of entropy rate is not limited to random processes, but it can also be defined in the case of deterministic dynamical systems. Let *x*_*t*_ be a continuous univariate times series. Then we can construct a state-space representation of the process as in Equation 2.12, if we choose an appropriate dimension *d* of the presumed underlying dynamical system and an embedding delay *τ* [23; 24].

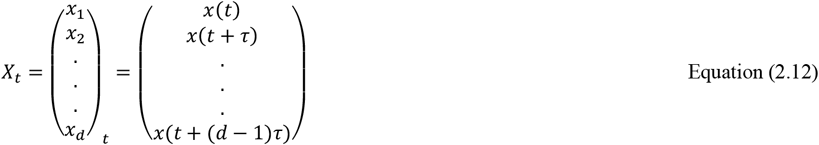

The existence and calculations of the embedding dimension and delay are ensured by Taken’s embedding theorem [25]. For more information on dynamical systems theory see *e.g*., [26; 27]. Essentially, any univariate time series can be viewed as being sampled from a high dimensional dynamical system [28]. The dynamical system follows a trajectory in the d-dimensional space defined by the *d* degrees of freedom. All possible states of the dynamical system define the phase space of the system.

If we partition the phase space across *F* dimensions, with *F* ≤ *d*, we have an F-dimensional grid of cells of volume *N*^*F*^. Then, we can measure the state of the system at constant time intervals equal to the embdedding delay *τ*. Then we can define the joint probability *p*(*i*_1_, *i*_2_, …, *i*_*d*_) that *X*_*τ*_ is in cell *i*_1_, *X*_2*τ*_ is in cell *i*_2_,…., *X*_*dτ*_ is in cell *i*_*d*_. The degree of “randomness” of the determistic system can then be calculated using the Kolmogorov-Sinai (KS) entropy[29] using Equation 2.13:

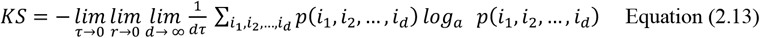

The KS entropy is almost always equal to the entropy rate of the original signal *x*_*t*_ and characterizes the degree of randomness of the system (and subsequently the sampled one-dimensional signal). For completely deterministic systems it is equal to zero and it is infinite for random systems.

In practice, the entropy rate is approximated using what is known as the correlation integral [30] in Equation 2.14:

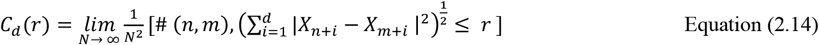

*i.e*., the (#) number of pairs of trajectory points that are close to each within a tolerance threshold *N* and measures the regularity (frequency) of patterns like a given template of specific length.

It can be shown that:

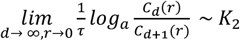

Where *K*_2_ is the Renyi entropy of order 2. The Renyi entropy *K*_*P*_ in Equation 2.15 is a generalized form of the usual Shannon entropy and is defined as:

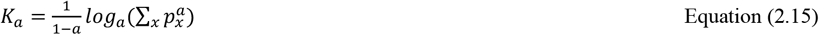

We leverage these tools to calculate the entropy rate in the case of a discrete time series *u*(*n*). Consider two different blocks of length *m* sampled from the time series:

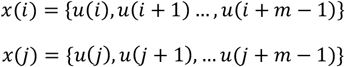

And define the distance in Equation 2.16:

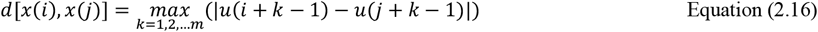

*i.e*., the maximum distance between the two vectors (Chebyshev distance). Then, we can define a quantity in Equation 2.17 like the correlation integral, for a template of length *N* at *x*(*i*) within a tolerance threshold r:

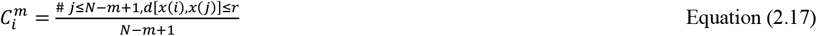

Then, the entropy rate can be estimated as in Equation 2.18:

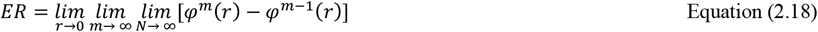

Where as in [31] Equation 2.19 gives:

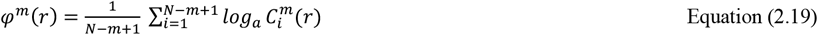

Since 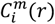 is essentially the probability that any sequence of length *N* is very close to the template sequence at time *i*, and 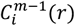 the probability that the same holds true for sequences of length *m* − 1, then 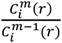 is the conditional probability that any sequence of length *N* is very close to the template of length *m* at time *i* given that the same holds true for *m* − 1. Then 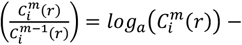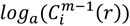 the logarithm of this conditional probability. It is easy to see that *φ*^*m*^(*r*) − *φ*^*m*−1^(*r*) is the average over *i* of the logarithm of this conditional probability [29].

However, due to finite sample sizes and stochasticity in time series analysis, the entropy rate can be estimated by what is known as Approximate Entropy [26] and is given by Equation 2.20:

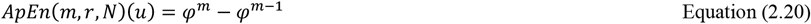

Where *N* is the length of the time series *u*(*n*), *N* is the choice of the length template and *N* is the threshold tolerance choice. Approximate entropy measures the logarithmic frequency with which segments of length *N* that very close together (according to the threshold), stay together through time.

An approximate formula for ApEn, which we implemented in our study is given by Equation 2.21:

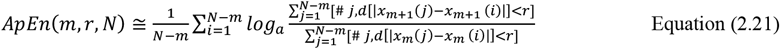

### 2.10 Entropy Rate estimation for a binary MMS speed sequence

Generally, in the case of discrete alphabet sequences with *N* symbols, 0 ≤ *ApEn* ≤ *log*_*P*_ *N*

Where *ApEn =* 0 for deterministic time series and *ApEn = log*_*a*_ *N* for random series. In our case (binary MM series), *N =* 2 and 0 ≤ *ApEn*_*MM*_ ≤ *log*_*P*_ 2

For a *=* e (natural logarithm choice), the maximum value is *ln*(2) *=* 0.69, which the base we use in this study [29].

A good choice of *N* is equal to the embedding dimension, which can be estimated using the False Nearest Neighbor (FNN) algorithm[32]. Usually, *N* is of low dimension, in our case the dimension of the data was estimated to be 2. The threshold *N* is usually set between 0.1 to 0.25 standard deviations of the time series [29].

### 2.11 Quantifying Information Flow Between Binarized MMS with Local Transfer Entropy

Local Shannon Entropy is defined in Equation 2.22 as the negative logarithm of the probability of an outcome *x* of a random variable [33]:

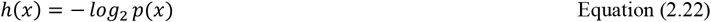

where low probability outcomes carry more information than high probability outcomes. Entropy as defined in Equation 2.23 can then be expressed as the average value of all such outcomes:

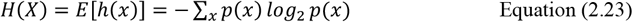

Where *E*[.] is the expectation (average) operator. An estimator based on samples *x*_*n*_ is given by Equation 2.24:

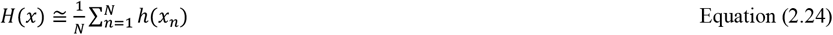

The Local Mutual Information *i*(*x*; *y*) and Mutual Information (MI) *I*(*X*; *Y*) are respectively defined in Equations 2.25 and 2.26, [34]:

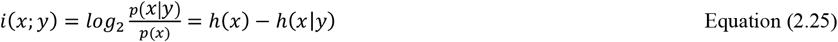

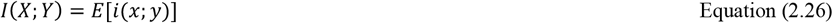

Equation 2.26 quantifies the information that we gain when observing *X* after we have already observed another variable *Y*.

Similarly, the Local Conditional Mutual Information and Conditional Mutual Information are given by Equations 2.27 and 2.28 respectively, [34]:

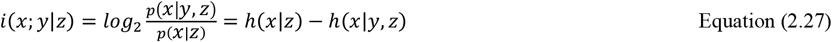

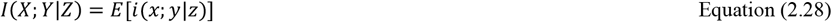

It quantifies the information that we gain when we observe *X* after considering both *Y* and *Z* versus considering only *Z*.

Finally, local transfer entropy quantifies the flow of information from Y to X and is defined in Equation 2.29, [35; 36]:

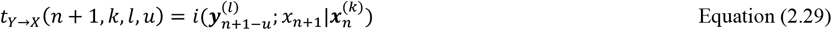

Where *l* and *N* denote the length of the vectors 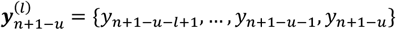 (storing past information of the process Y with a memory of *l* samples up to point *n* + 1 − *u*) and 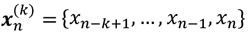.

The integer *u* denotes the source-destination lag, *i.e*., the causal time delay between *Y* and *X* that we are interested in when we want to calculate the transfer entropy from *Y* to *X*. For *u =* 1, a typical choice of source-destination lag is given by Equation 2.30:

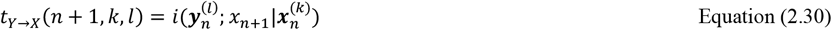

The local transfer entropy is the mutual information between Y and the future state of X, *u* samples ahead, conditioned on the history of X. In other words, it measures the information gained that we get about the future state of X when considering both its own past and the past states of Y versus considering only its past state. Transfer entropy is the expected information gain, averaging over all states given by Equation 2.31, [33; 35]:

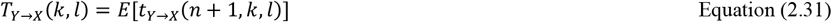

### 2.12 Quantifying Autonomy of an Agent from the Perspective of the Observer

For a child/clinician dyad, we obtain the normalized MMS derived from the fluctuations in angular speed from the right- and left-wrist sensors throughout the course of the dyadic interaction. Then, we calculate the entropy rate for consecutive non-overlapping time windows, small enough to ensure stationarity but not too small, as to ensure convergence. We calculate the entropy rate both for the normalized MMS and for the corresponding binary MMS trains that we obtain by setting peak values to “1” and zero values to “zero”.

To estimate the entropy rate we used Approximate entropy ApEn (developed by Steve M. Pincus [37]), which measures the amount of regularity or unpredictability of fluctuations over time-series data that have lengths compatible with experimental settings (unlike other measures of entropy aimed at measuring regularity but requiring very long times). There are caveats to the use of the ApEn algorithm [29]:

i. The ApEn algorithms allows self-counting when counting the number of templates that are similar to a given data segment, which helps avoid the occurrence of log(0) in the calculation.
ii. However, when the self-similarity threshold *N* is very small, the template vector coincides only with itself, giving ApEn low values, indicating regularity when the system may in fact, be very irregular.
iii. ApEn is biased by a factor of 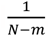, which means that it depends on the template length and data stream length.

ApEn generally depends on the threshold *N*, and the embedding delay and embedding dimension of the reconstructed space (which is equal to the template length). It is generally suggested, that in order to compare the approximate entropies of different time series, all parameters must be equal. However, for the scope of our study, we chose the threshold parameter *N* to be equal to 0.2*σ*, as recommended in the literature [29]. The embedding delay was chosen according to the minimum Average Mutual Information criterion, to ensure maximum novelty between consecutive samples in the reconstructed space. As for the template, we chose it to be 1/*N*, where *N* is the average rate (frequency) of MMS in the time window of interest. This equals to the average time-distance between two spikes and our choice ensures that in the reconstructed space, the coordinates of a point in time include both zeros (“*quiet moments*”) and spikes and that the system does not bounce back and forth from a single coordinate of zeros components. In this way, we can minimize any bias introduced by differences in spike rates, in the computation of self-similarity by the algorithm. Sparser time windows will contain the same percentage of “*active*” moments as denser time windows. Since it turns out that, for our datasets, 0.1 < R < 0.5, we have 2 < *m* < 10, which according to the literature is within the optimal range [29]. Moreover, since *N =* 1000, the bias introduced by *m* in the prefactor is very small.

ApEn is computationally efficient. One can easily see that the worst-case time complexity of ApEn is *O*(*N*^2^). Furthermore, it has lower effect from noise in the data. If data is noisy, the ApEn measure can be compared to the noise level in the data to determine what quality of true information may be present in the data [29]. We here notice the difference between the criterion for randomness in the Gamma parameter space, when the shape is 1, which is the special case of the memoryless exponential distribution. In our empirical characterization of the MMS from the peak fluctuations, which follow a scaling power law, as the shape approaches the value of 1 representing the exponential distribution case, the *NSR = logθ* approaches its maximum levels [3]. The differential entropy for the Gamma distribution has the general form in Equation 2.32, [38]:

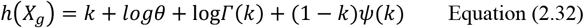

We will show later that discrete samples *X*_*G*_ that follow the Gamma distribution, such as the MMS, have entropy roughly equal to *h*(*X*)_*g*_ − *logΔ*, when *Δ*, is the discretization step. Because of the Power Law discussed before, *log*(*k*) = *a* + *b log*(*θθ*) + *ε*, we have in Equation 2.33:

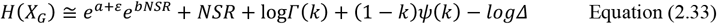

As the NSR increases, *k* → 1 and thus, 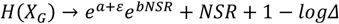. In Section 4, we will experimentally show that, for *logθ* < 1 in Equation 2.34:

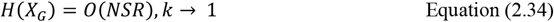

However, in the case of ApEn, we consider a process of the form 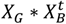, where 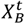 is the binary spike series, determining the temporal distribution of the peaks in time. In fact, we will empirically demonstrate that 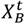 is almost independent from *X*_*G*_, implying that ApEn measures the information content of the binary spike time series, characterizing the motor code. On the other hand, randomness in the sense of *NSR* (or equivalently *H*(*X*_*G*_)), refers to the temporal component of the events and answers the question of predictability in time, whereby predicting future events in time does not benefit from knowledge of prior or current event times. We will see later that these two elements of the Gamma distributed MMS are indeed separable and within the current context, tend to be orthogonal.

In this sense, we propose that the entropy rate (ER) derived from ApEn is a measure to characterize autonomy in the system. Since ER is a proper way to quantify regularity *vs*. randomness, we can safely presume that the information levels that it carries also measures the ability of an observer to predict the motor behavior of an agent, when the two of them engage in a dyadic social interaction. For example, when the clinician observes the behavior of a child that engages in repetitive and predictable motions, they can easily learn their behavioral and motor patterns. This also implies that they can more easily detect a problem in a child that behaves predictably and set up the context to better control the situation. In this sense, the more predictable the situation is, the more control it will be afforded by the external agent.

Following our argument, we redefine socio-motor agency as the balance between control and autonomy. Signal-to-Noise ratio characterizes the ability of an agent to (internally) control their own behavior. Entropy rate characterizes the ability of the agent to act autonomously (while minimizing external control by another agent) in a social interaction.

Finally, using transfer entropy we can quantify the amount of causal influence from the clinician to the child and vice versa, without the need to use any model or make any other assumptions.

**Methods Figure.**
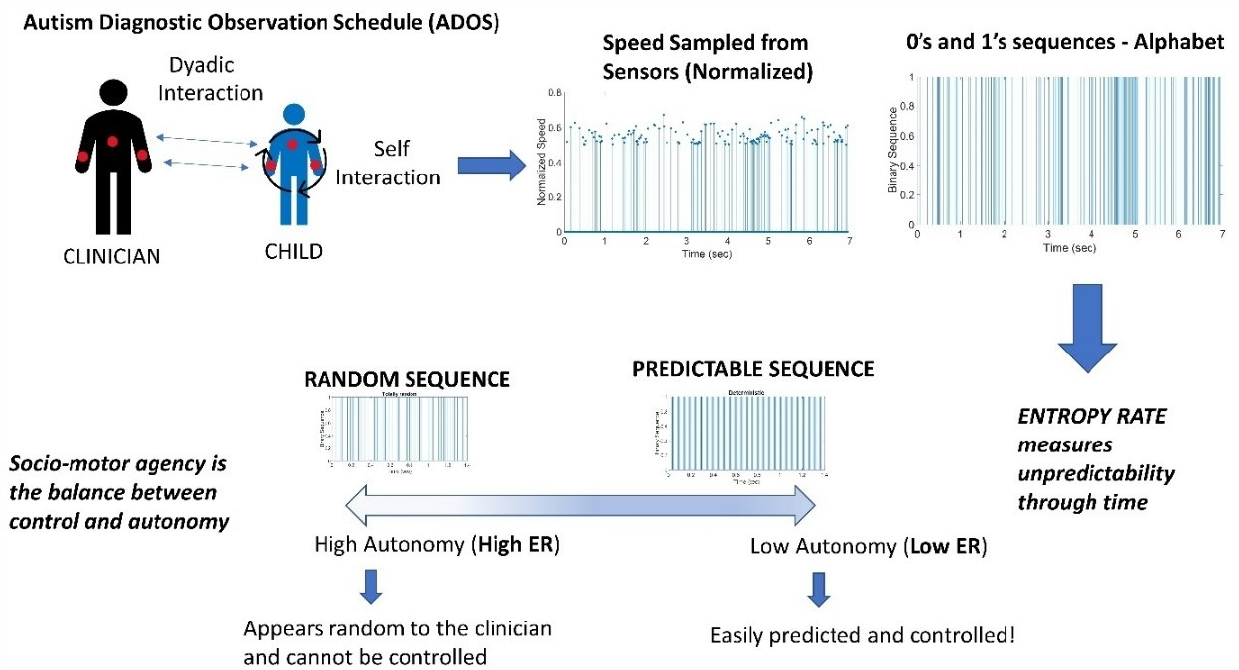
Digitization of the Autism Diagnostic Observation Schedule (ADOS): Angular speed samples (128 Hz) from wearable sensors on the wrists and torso of the child and clinician are normalized and binarized to obtain discrete sequences of 0’s and 1’s. Entropy rate estimates measure the unpredictability of the underlying binary processes to characterize the agents’ autonomy in the dyadic social interaction. The analysis is performed on data from time windows of ∼7.8 secs which proved optimal to attain high confidence intervals.

### 2.13 The Autism Diagnostic Observation Schedule (ADOS-2) Scoring System

The ADOS-2 Modules consist of tasks that the clinician performs with the child to observe behavior related to the diagnosis of ASD and reach a conclusion. There are different Modules. Each child is administered a single Module based on their expressive language level, developmental age and their unique interests and abilities. However, they are designed in such a way that ensures that judgements about social and communicative abilities are as independent as possible from level of language ability and chronological age.

Both Modules (toddler) T and 1 are administrated to non-speaking children, Module T for ages 12-30 months and Module 1 for children over 31 months. Module 2 is administrated to children of all ages who are using phrase speech but are not yet speaking verbally with fluency. Modules 3 and 4 are administrated to individuals that are speaking with verbal fluency, with Module 3 specifically designed for Children / Younger adolescents that can still play with action figure-type toys and Module 4 for older adults. All Modules are administrated under the assumption that the individual can walk independently and is free of visual or hearing impairments. This assumption is erroneous, but we use the ADOS-2 test not to diagnose but to evoke social situations leading to movement patterns likely present in such situations.

Our current analysis focuses on Modules 1,3 and 4. We suggest the following categorization of tasks to better relate our digital biomarkers to the clinical tasks that evoke some aspect of social interactions and emotions present in human gestural communication, which is mediated by movements:

#### Socio-Motor Tasks

These are tasks that engage interactive movements within the child, the clinician, and jointly between the child and clinician. Construction Task, Joint Interactive Play, Demonstration Task, Cartoons, Conversation and Reporting and Break Tasks all have in common the Child’s Socio-Motor behavior involvement. Construction Task consists of an interaction between the Clinician and Child that involves reaching over the Clinician’s arm to ask for block pieces that may form a shape. Joint Interactive Play consists of a Play Sequence between the Child and the Clinician that involves body movements. During Demonstration Task the Child uses their body to represent objects and mime the use of each object. During Cartoon Task, the Clinician observes the Child’s gestures and coordination with speech. Similarly, during Conversation and Reporting body language and facial expressions / gestures are observed alongside general communicative skills. During Break the Child is expected to move around the room.

#### Emotional Tasks

These are tasks that probe the child’s emotional states. Emotions, Social Difficulties, Friends, Relationships, and Marriage and Loneliness all evoke strong emotional responses from the Child. During the Emotions Task, the Child is asked questions about social relationships, different emotions such as happiness, fear and anxiety and details about the manifestation of these emotions under different circumstances. Social Difficulties and Annoyance consist of questions related to social interactions at school or work, such as bullying or teasing. Friends, Relationships, and Marriage are designed to evaluate the Child’s concepts on topics such as friendship and social relationships and the questions asked can cause strong emotions in the Child. Similarly, during Loneliness task, questions are asked about the concept of loneliness, which is a heavy topic, especially for Children on the Autism Spectrum, that struggle with social rejection and bullying from a young age.

#### Abstract Tasks

These are tasks that require higher, abstract-level of cognition. Make-Believe Play, Description of a Picture, Telling Story from a Book, and Creating a Story all help observe higher cognitive skills. Make-Believe Play involves the use of dolls an action figures and the Child is tested for their ability to perceive them as animate beings and produce imaginative sequences of actions that involve these objects. Perception or the lack of it of objects as animate beings is a concept frequently encountered within the context of the Theory of Mind. During Description of a Picture Task the Clinician observes the Child’s use of language/ communication and the level of interest in the picture presented. Telling a Story from a Book is similar but involves a story from a book instead of a picture and humor and presumption of the feelings of the characters from the book are evaluated as well.

After the administration of Module 3 a scoring system is used to evaluate the levels of Social Affect (SA) and Restricted and Repetitive Behavior (RRB). The scores are added up to determine a final score, from 0 to 10. A score of 0 or 1 indicates Minimal to No Evidence of ASD related symptoms, scores between 2 and 4 indicate a Low Level of ASD related symptoms, 5 to 7 Moderate and 8 to 10 High. A score of 9 or more determine that the Child is Autistic whereas a score of 7 of more that the Child is in the Autism Spectrum. Furthermore, Social Affect consist of Communication (Reporting of Events, Conversation and Descriptive, Conventional, Instrumental, or Informational Gestures) and Reciprocal Social Interaction (Unusual Eye Contact, Facial Expression Directed to Examiner, Shared Enjoyment in Interaction, Quality of Social Overtures, Amount of Reciprocal Social Communication, Overall Quality of Rapport) Scorings. RRB consists of scoring Stereotyped/ Idiosyncratic Use of Words or Phrases, Unusual Sensory Interest in Play Material/ Person, Hand and Finger and Other Complex Mannerisms and Excessive Interest or Highly Specific Topics/ Objects or Repetitive Behaviors.

## 3 Results

### 3.1 Age-dependent Dyadic Motor Control Separates Neurotypical (NT) from Children on the Autism Spectrum Disorders (ASD)

The micro-movement spikes (MMS) derived from the biosensors’ signals, offer a standardized time series that scales out anatomical differences across participating children of diverse ages. This standardized signal within the ADOS-2 tasks contexts, is well characterized by a continuous Gamma process. Here (as in prior work involving other biosensors) the Gamma shape ***k*** and the Gamma scale ***θ*** parameters can be empirically estimated from the normalized spikes, using Maximum Likelihood Estimation (MLE) with 95% confidence [3; 39]. The normalized spikes, which conserve the original time latencies of the raw peaks, represent a time series of quiet times (at averaged activity) interspersed with bouts of activity evoked individually for each child and pertinent to the task at hand. We measure these biorhythms individually for the child’s and clinician’s dominant hand. We also measure them from the shared, synchronous activities of the social dyad composed by the child and the clinician.

The empirically estimated Gamma parameters localize each child-clinician’s dyadic interaction for each task on the Gamma parameter plane with 95% confidence intervals (Figure 2A). These points represent the empirical probability density function (PDF) of their joint dyadic interaction. When we plot the full scatter estimated from each task in the ADOS, for all children, a tight linear relation emerges whereby the log-log plot follows a scaling power law, of the form ***k*** ≅ ***aθ***^***b***^. (See methods for a more in-depth analysis of the power law and the micro-movement spikes (MMS) time series transformation).

**Figure 2.**
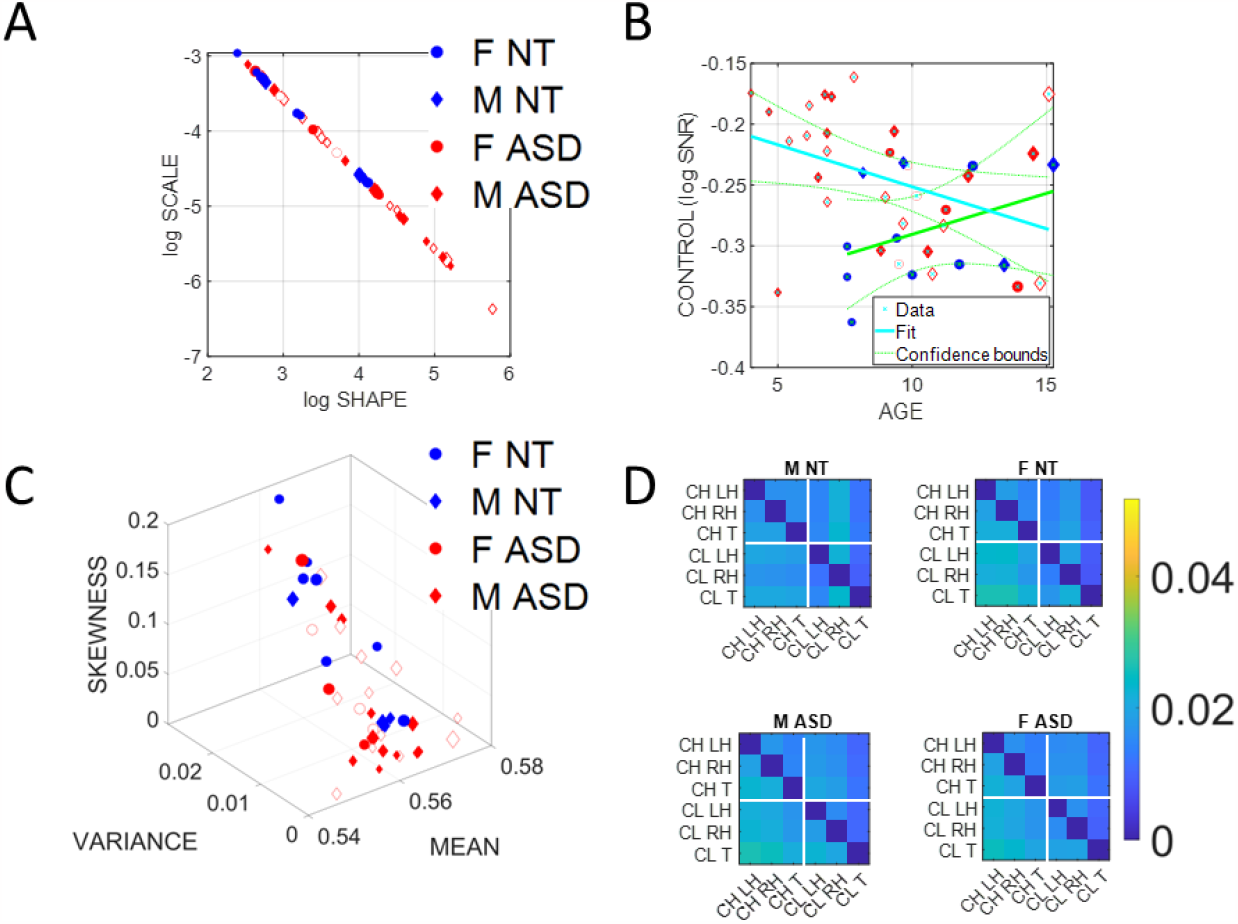
Summary stochastic characterization of micro-movement spikes, MMS, derived from ADOS-driven dyadic interactions, using angular speed registered from the right (dominant) wrist. Activity encompasses the entire ADOS session. Filled markers represent first visits to the clinician, unfilled markers are subsequent visits. (A) Empirically estimated Gamma Shape and Scale (NSR) for each participant using Modules 1, 3 and 4 of the ADOS test as a backdrop behavioral assay. The size of the marker is proportional to the age of the participants. Empirical Gamma plane of individual children and clinicians separating young from older children and adults in an interpretable map of human neuromotor maturation. (B) Child’s negative Gamma scale parameter (log -NSR = log SNR) denotes control as a function of age. Observe (cyan line) the decreasing trend of SNR with age for ASD in contrast to the opposite trend for NT (green line). (C) Parameter space spanned by the empirically estimated Gamma mean (x-axis), standard deviation (y-axis) and skewness (z-axis) derived in (A). Marker size is proportional to age. (D) Quantification of Transfer Entropy for social dyads involving clinician and child obtained for males and females in the NT and the ASD groups, using 6 sensors, 3 on the clinician and 3 on the child, outputting time series of angular speed motion on the left and right wrists and the trunk of each social agent in the dyad. Off-diagonal entries represent joint dyadic activities.

This relationship, first described as a maturation law in humans’ voluntary decision-making, mediated by pointing motions [3], is reproduced here for gyroscopic data reflecting joint dyadic angular speed, such that as the Gamma scale value decreases, the Gamma shape value increases. Because knowing one, we can predict the other with high certainty, we can then reduce these two parameters of interest to one parameter summarizing these motor signatures of the interacting dyad. We can also do so for each individual signature, *i.e*., those of the child and those of the clinician in standalone mode.

Importantly, the continuous Gamma family of probability distributions has the first (mean) and second (variance) moments expressed in terms of the shape and scale as in Equations 3.1:

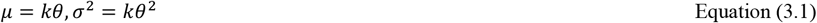

Then, unfolding Equation 3.1, the Noise-to-Signal Ratio (NSR) of the MMS reduces to:

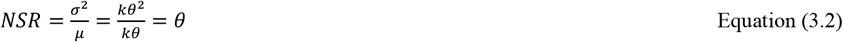

The Gamma scale parameter in Equation 3.2 fully characterizes the noise of the biorhythmic motor patterns of the interactive dyad, *i.e*., in relation to their joint level of fluctuations of angular speed during the ADOS activities.

Empirical estimation of these parameters across thousands of participants over a decade of work involving humans along the lifespan, and across disorders of the nervous system, in voluntary, involuntary, spontaneous, and autonomic processes, has revealed an interpretation for the Gamma log-log parameter plane. Distributions that fall along high NSR regimes are also close to the memoryless random regime of the special exponential distribution case (when *k = 1*). Points in mid NSR correspond to heavy-tailed Gamma distributions. Then, low NSR (or high signal = 1/NSR) are congruent with symmetric shapes (Gaussian-like) distributions.

High-signal Gaussian regimens are highly predictable in contrast to High-noise memoryless random Exponential regimes. As such, the Gamma parameter plane is empirically interpretable and the locations of the distributions representing the shape and scale signatures of individual participants change in an orderly ontogenetic manner whereby a decrease in the NSR is accompanied by a drecrease in randomness (away from the memoryless exponential distribution at shape = 1). The lack of maturation of the human nervous system is well characterized by high NSR and random fluctuations previously found across ASD [3]. In this sense, we equate high SNR=1/NSR with an index of controllability. As per the scaling power law, high SNR of the MMS is equated with high predictability of the person’s self-referenced, self-generated motor code. This motor code represents a proxy of kinesthetic reafference, *i.e*., the continuous stream of motor activity from the periphery, serving as an index reflecting the quality of the motor feedback to the central controller of the nervous system.

Then, as this motor code is shared with another agent during social dyadic interactions, the distributions of the joint dyadic interactions of the participant and the clinician, for the 26 participants (11 neurotypically developing NT and 15 ASD), can be appreciated in Figure 2A following a power law. These distributions are derived from the MMS that fluctuations in angular speed produced in the dominant hand (see Methods Figure).

Furthermore, Figure 2B shows that the log(SNR)=-log(NSR) (denoted as the index of control) of the interacting socio-motor dyad has an age-dependent pattern. In NT children, as the age increases, control tends to slightly increase *i.e*., a slight positive trend is reflected in the slope of the line fitting the (blue

NT scatter), NT: intercept = 3.0271 p = 0.0067, slope = 0.0786, p = 0.362. In contrast, as ASD children age, control tends to decrease, *i.e*., a strong negative trend is quantified in the slope of the line best fitting the red ASD scatter, intercept = 5.5317 p = 5.65 x 10^−12^, slope = -0.1074, p = 0.0463.

In Figure 2C we compare the two groups by localizing each participant on the Gamma moments space spanned by the empirical mean, variance, and skewness, whereby each point represents the empirically estimated moments of the Gamma PDF of joint dyadic activity for each child-clinician pair. This result demonstrates a tendency of the joint dyad moments to separate NTs from ASD participants, as they interact with an adult clinician, expressing marked differences between males and females.

To better appreciate the sex differences, we obtain pairwise the Transfer Entropy (TE) from child to clinician and from clinician to child. See methods for an extended definition, but recall that TE is the reduction in uncertainty of predicting the future of X when we consider the process Y. In Figure 2D, we can see for each matrix the pattern that emerges when considering the time series data from each of the 6 sensors attached to the child’s and clinician’s two hands and trunk. The cross terms in the off-diagonal entries of the matrix (top right-hand entries 1,4 to 1,6; 2,4 to 2,6; 3,4 to 3,6; and bottom left-hand entries 4,1 to 6,1; 4,2 to 6,2 and 4,3 to 6,3) represent the dyadic cases of child → clinician and clinician → child, respectively. There, in the shared entries of the matrix we see that in ASD, males show a decrease in TE values while females show an increase. In the context of the ADOS, females evoke a reduction in the clinician’s uncertainty predicting the impending females’ motions, *i.e*., perhaps an inherent bias that partly accounts for the disparate ratio of 4-5 males per each female diagnosed with ASD. We will further explore these differences to try and understand the interplay between the NSR as an index of controllability (predictability) and the overall sense of socio-motor agency in each of the ADOS tasks, for males and for females.

In the diagonal sub-matrices (top left-hand entries 1,1 to 1,3; 2,1 to 2,3; 3,1 to 3,3; and bottom right-hand entries 4,4 to 4,6; 5,4 to 5,6 and 6,4 to 6,6) we represent the patterns within the individual’s body parts. There we appreciate higher values of TE from child → child in both ASD males and females, with ASD females having higher TE than ASD males. As with the shared dyadic activity, here in the individual patters, the highest differences for clinician → clinician can be appreciated in the ASD females.

### 3.2 Quantifying Dyadic Social Agency Reveals Differences Between NT and ASD

High levels of NSR in the MMS fluctuations from the angular speed coincide with memoryless random regimes of motor patterns - well characterized by the exponential distribution previously found in autistic individuals [3; 40]. It has been proposed that under such random and noisy motor code, it is difficult to have high quality motor feedback contributing to a predictive code. Such predictive code would be necessary to compensate for sensory-motor and inertial time delays inherent in the nervous system [3; 14; 40; 41; 42].

In a dynamic dyadic social interaction such as that taking place during the ADOS, it is then difficult to exert control over the interaction because presses by the clinician and overtures by the child are not occurring at the expected timely rates. This temporal mismatch in autism alone can bias the rating by the clinician in ways that differ between NT and ASD, but also differ between males and females. Here we equate high NSR with low predictive control and posit that the type of socio-motor agency required in a naturalistic social interaction will be impacted by poor controllability levels on one side of the dyad. We then question whether dyadic-based control (*i.e*., shared by the child and clinician) is differentially impacted in ASD participants.

Another aspect of dyadic social agency is autonomy. As mentioned earlier, autonomy is the ability of the child to lead the conversation as much as the clinician does, rather than always following the lead of the clinician. An obvious way to quantify the degree to which the clinician is leading would be by using some form of causal analysis between data recorded on the clinician and data recorded on the child. As our main approach however, we choose to quantify autonomy in ways that depend on data recorded from wearable sensors on a single agent which as we will show, is intuitive and can be applied in a clinical setting, to help digitize the ADOS.

We introduce (behavioral) spike trains from the MMS derived from the time series of angular speed. We use entropy metrics to examine the degree to which the spikes behave randomly or deterministically (*i.e*., containing periodic, systematic patterns.) To that end, we use entropy rate, a metric well suited to interrogate the stochastic regimes of spike trains [26; 27].

From the MMS derived from time series of angular speed, recorded either from the left or the right hand of the child, we derive binary sequences whereby a sequence of 1’s corresponds to sudden bouts of activity and 0’s to “quiet” sampling periods, when no significant change above the person’s average activity occurs in the angular speed profile. Another way to view these binary sequences is as the manifestation of an underlying “alphabet” that characterizes the predictability of the motor code. Zeros and ones will appear with some probability, which we expect to change at some time scale, due to the non-stationary nature of human motion. But if we restrict ourselves to small time windows (∼7.8 seconds, determined as optimal for empirically estimated confidence intervals, upon sampling different sizes), this time widow is small enough that the process can be viewed as stationary, yet large enough to contain a satisfactory number of samples lending statistical power to our empirical estimation per window. As such, 7.8 seconds is our unit of time for the spike trains that we derived. Using this MMS per unit of time as our data type, we can then measure the degree of randomness of the child’s motions, by estimating the entropy rate (see Methods). Furthermore, we then compare it to transfer entropy (TE) obtained from the child and clinician, a causal metric that can inform us of who leads the interaction for any given task.

We argue that a suitable scale of autonomy is one in which, at one extreme, a high degree of randomness is a measure of a system at its highest degree of autonomy. This is the type of state where the system is uncontrollably “hidden” from the controller. There is no opportunity to control the person.

At the other end, the lowest degree of randomness leads to a systematic, deterministic pattern, highly controllable. While in the former, the child’s system with excessive autonomy prevents social exchange with the clinician in that the clinician cannot control the child, in the latter, the clinician can absolutely control the child. Either extreme is detrimental to the development of rapport or turn-taking in a social exchange. A happy medium is one in which while the child preserves a degree of autonomy that enables a balanced social exchange, the clinician also partakes in a give-and-take interaction, rather than leading the child most of the time.

We test our new hypothesis that motor autonomy relates to measures of entropy by comparing TE (a measure of causality) from the child to the clinician, with entropy rate, a measure spanning a scale from totally random to totally deterministic states of the spike-based code. We show in Figure 3A an age-dependent trend spanning two scatters. In older neurotypical children, the scatter aligns such that as the child’s entropy rate (denoting a scale of autonomy) increases, so does the TE denoting a causal lead of the child over the clinician. In contrast a second scatter emerges for younger children whereby the trend is less visible, indicating that these children’s index of autonomy is not as evident during the exchange and the causal lead (TE) denoting the child’s lead over the clinician’s lead, is less evident.

**Figure 3.**
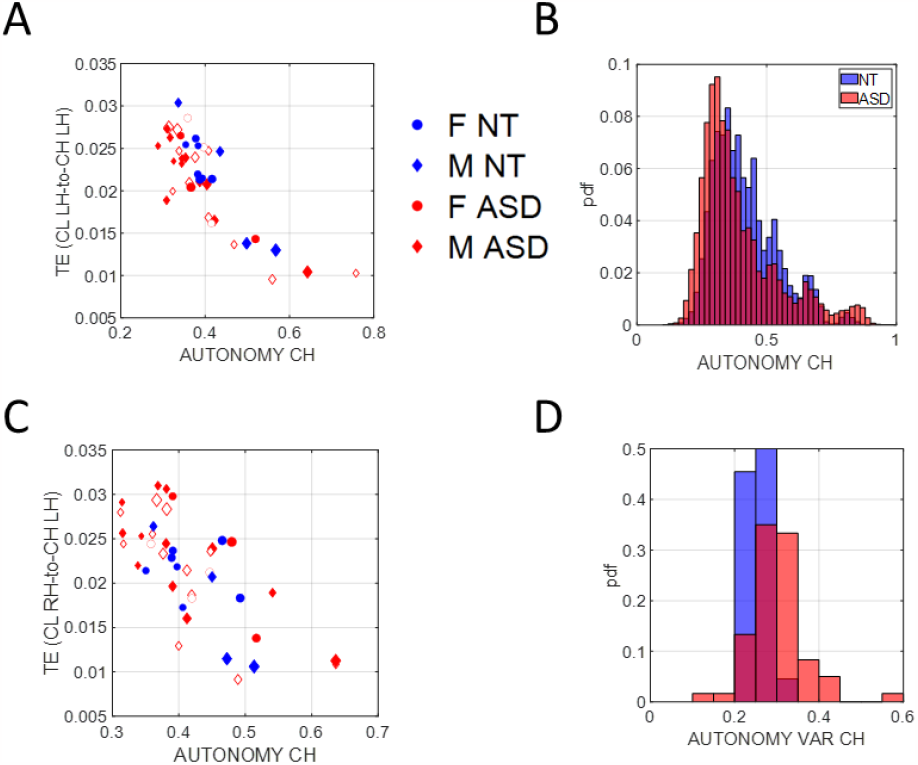
Scales of socio-motor agency according to index of autonomy. (A) Average transfer entropies between child→clinician taken over windows of ∼7.8 secs duration, per participant (filled in markers are first visit to the clinician, unfilled markers are subsequent visits) vs. autonomy (Ap Ent) reveal higher autonomy index in NT, a trend that is also quantified in (B). As the child autonomy decreases, the CL→CH TE (left hand) decreases. Adding the CL past activity does not contribute more information about the CH state than looking at the CH past activity alone. (C) This is also the case for the right hand. (D) Autonomy variability (variance over the mean) throughout a session, is higher for the ASD group, both for the child and the clinician involved.

We can appreciate the shift in this metric of autonomy in Figure 3B where the histogram of the ASD children is shifted to the left, indicating lower density values than NT children.

Since the left hand is not the dominant hand in these children, we plotted the histograms pertaining to the left hand as well, to see if these effects consistently emerged. We see in Figure 3C that across multiple time windows, the pdf for the neurotypical group is shifted to the right, meaning that on average, NTs have higher values of autonomy than ASDs. Since autonomy also varies throughout sessions, plotting the autonomy variability (variance of this index over the mean of this index) for different participants in Figure 3D shows that for ASDs, child and clinician variability is higher than most NTs. This variability index tends to separate ASD from NT participants, particularly for later visits (as the child aged, over 2 years and a half that the study spanned.)

### 3.3 Age-Dependent Autonomy Across Children *vs*. Clinician’s Autonomy Robustness

As we saw earlier, the SNR (1/NSR) of the control index, has trend with age that differs between the two groups. NT children show increasing control with age, whereas ASD children show a decreasing trend. Likewise, here we ask if the index of autonomy also changes with age. To that end, we examine this index as a function of age across the children. We also examine it for the clinician across the children’s ages.

We find that the child’s index of autonomy for both NT and ASD increases with age in all cases (Figure 4A). This result reveals that the ability of the ASD child to actively participate in a dyadic interaction is a human socio-motor developmental trait that improves with age. In contrast, Figure 4B shows that the clinician’s autonomy is independent of the child’s age. In this case, the adult clinician shows no discernable trend.

**Figure 4.**
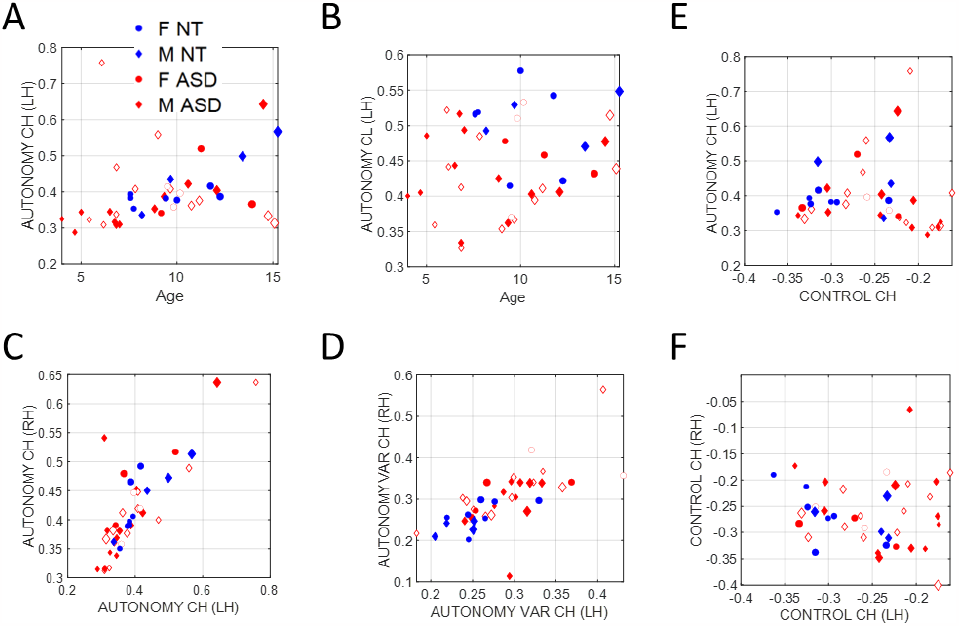
Non-equivalence of the index of autonomy and the index of control. Plots reflect the average child and clinician index of autonomy for left hand motions *vs*. age as well as right *vs*. left hand index of autonomy, index of autonomy variability and index of control. (A) Child index of autonomy is positively and linearly correlated with age. (B) There is no trend between the clinician index of autonomy across children’s ages. (C) Equivalence of index of autonomy derived from the left hand *vs*. the right-hand motions. (D) Index of autonomy variability also correlates between the two hands and separates NT (blue) *vs*. ASD (red). (E) No definite relationship between index of control and index of autonomy is observed, however for small values of control index there seems to be a positive trend which then becomes negative for high values. (F) Left hand motions have higher variability in the index of control than do right hand motions.

### 3.4 Indexes of Autonomy and Control are Not Equivalent

The child’s index of autonomy and the variability of this index extracted from the sensors in both hands, are linearly correlated (Figure 4C). In the case of the index of control however, there is higher variability of the mean autonomy index across subjects when we use data from the left-hand sensor (as shown in Figure 4D, where separation of the NT from ASD is evident). For this reason, we focused our analysis on the non-dominant, left-hand motions. Furthermore, the index of autonomy derived from the left-hand motions as well as its index of control, are positively correlated for small values of control index, but negatively correlated for higher values. This is shown in Figure 4E. In other words, autonomy and control are not equivalent metrics. This can be further appreciated in Figures 4D (autonomy variability) *vs*. Figure 4F (index of control.)

### 3.5 Male vs. Females Respond Different to ADOS Tasks - The case of ASD Females

Besides the quantification of *indexes of* control and autonomy as components of socio-motor agency, we rendered important to consider the heterogeneity of tasks in the ADOS’ modules 1, 3 and 4 used here across children with different levels of spoken language. We grouped tasks into three main categories: Socio-Motor, requiring high motoric components (frequent movements and gestures); Abstract, tasks more “mental” in nature, requiring abstraction, theory of mind, and other cognitive components; Emotional, tasks that elicit feelings and emotional reactions, strongly *visibly* impacting the child’s emotional states.

We calculated the average indexes of autonomy and control across all participants, derived from samples corresponding to the different ADOS tasks. Then, we assessed potential differences between ASD and NT participants, focusing on the comparison of males *vs*. females. We found that ASD males respond with lower index of autonomy than do NT males. In contrast, ASD females *vs*. NT females, manifest very modest differences, inclusive of three tasks with no significant differences (Social Difficulties and Annoyance, Loneliness (both Emotional type tasks) and Construction Task (Socio-motor type task)).

It is therefore clear, that ADOS tasks inherently bear a lack of differentiation between NT and ASD females, unlike their male counterparts for which the differences are large. This can be appreciated in Figure 5AB (males) and Figure 5CD (females) where we color code the task type and code it numerically according to the name of the task (Methods describe the ADOS tasks included from each module.) Modest differences were observed between females. Tasks with nonsignificant differences in females were Social Difficulties and Annoyance, Loneliness (both Emotional type tasks) and Construction Task (Socio-motor type task). Notice that despite the non-significance, emotional tasks have broader spread in ASD females along the index of control than do NT females. In contrast, the index of autonomy has comparable spread for both. As socio-motor agency is defined as the ratio of index of autonomy/ index of control, this implies that across these ADOS emotional tasks, NT females have more social agency than ASD females. In contrast to females, a statistically significant difference between the two male groups was observed for all tasks. ASD males shift significantly to lower values of the index of autonomy across all tasks, but visibly socio-motor tasks are deeply affected.

**Figure 5.**
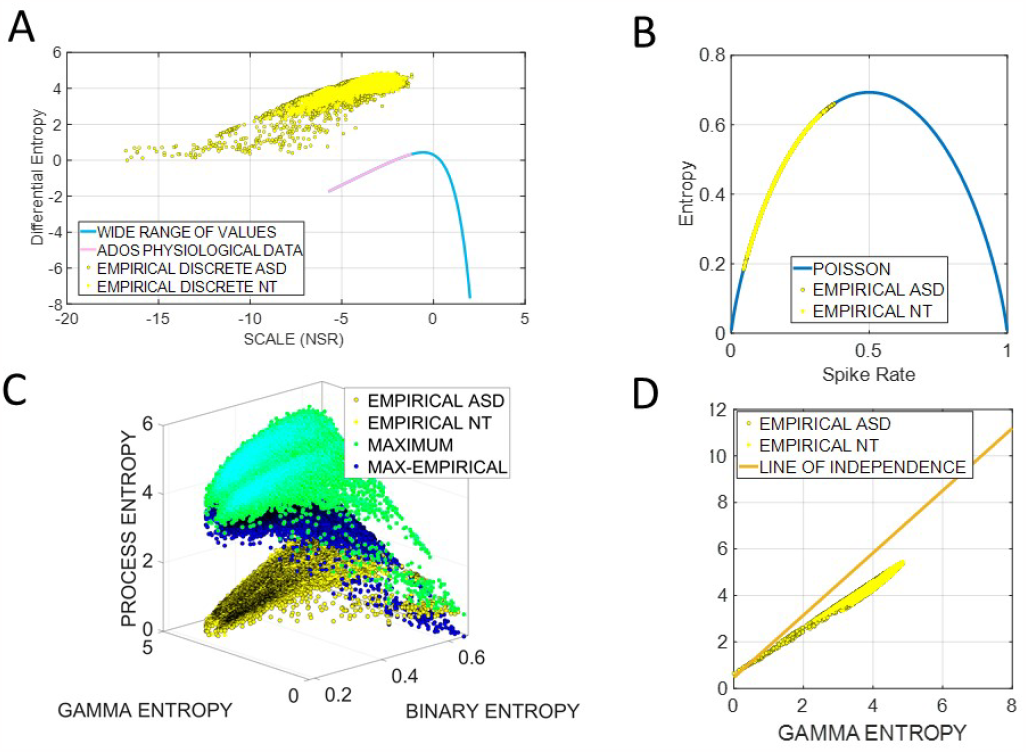
Differences between males and females in average index of autonomy *vs*. control. Filled circles code non-significant differences at the .05 level, while non-filled circles denote significant differences between NT and ASD participants. (A) NT females. (B) ASD females. (C) NT males. (D) ASD males.

## 4 Theoretical Considerations at the Intersection of Stochastic Analyses and Information Theoretical Metrics

The following section of the paper aims at exploring the relationship between the temporal code of the binary spikes embedded in the Gamma process, and the Gamma process itself, spanning information about the fluctuations in spike amplitude and inter-peak-timings. The latter follows a Poisson process, while the former is more generally revealing of multiple overlapping processes. Part of our quest is to try and deconvolve these overlapping processes in physiological data that contains multiple afferent streams from different levels of functionality. These levels may span from autonomic (pacemaker like regularities) to reflexive, to involuntary, to spontaneous and automatic, to voluntary levels of control previously proposed {Torres, 2011 #241}. At the core of our proposed measure of socio-motor agency lies the balance between bottom-up autonomy and top-down control, which we track through the spatio-temporal code of the time series of spikes.

### 4.1 Entropy-Spike rate and NSR relationship in the case of standardized biometric data sampled from child-clinician dyadic interactions

Recall that the entropy rate of a discrete process is defined as:

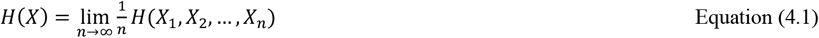

The case in Equation 4.2 is when all *X*_*i*_ are independently identically distributed *(i.i.d.)* [43]:

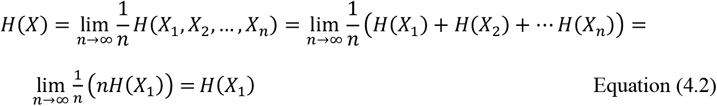

If *X*_*i*_ have identical entropies but are not independent, the following inequality holds in Equation 4.3 [43]:

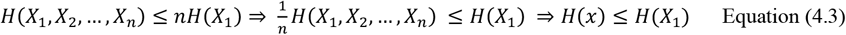

The standardized MMS processes that we extract from the wearable sensors consist of non-zero values (“peaks” that are gamma distributed) and zero values (“*quiet moments*” at the person’s average level of activity) and can be treated as the product between two processes. A gamma distributed process 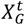 and a binary process 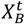, where *t* denotes discrete time. If for small time windows the process is roughly stationary, then at each point in time we have the processes *X*_*G*_ and *X*_*B*_, respectively. Then, from Equation (4.3) we have the upper bound for the entropy rate expressed in Equation 4.4:

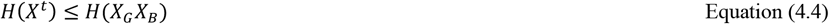

If *H*(*X*_*G*_, *X*_*B*_) is the joint entropy, because *f*(*x, y*) *= xy* is a measurable function, we have Equation 4.5 [43]:

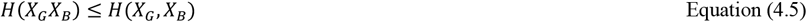

The upper bound for the joint entropy is expressed in Equation 4.6, [43]:

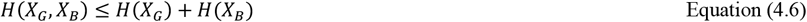

Equality holds true if and only if *X*_*G*_ and *X*_*B*_ are independent. Ultimately, from Equations (4.4) - (4.6):

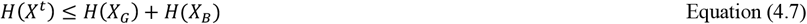

In practice, *X*_*G*_ is a discrete approximation of a continuous gamma variable *X*_*g*_. If Δ is the size of the bin used in the approximation and *h*(*X*_*g*_) the differential (continuous) entropy of *X*_*g*_, it can be shown that [43]:

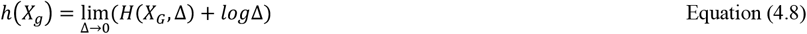

Therefore, for small Δ we can write:

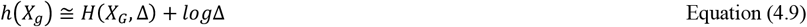

Equivalently:

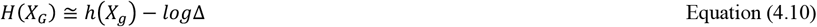

We mentioned earlier that the differential entropy of the gamma distribution (with shape *N* and scale *θθ*) has the closed form:

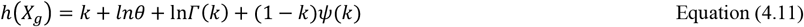

where *Γ*(*N*) is the gamma function and *ψ*(*N*) is the digamma function.

Because of the power law between shape and scale that we discovered from analyzing human motion data:

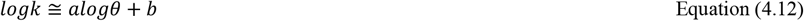

Where *N* ≤ 0 and b are parameters that are determined by fitting a regression model on the population of interest. Because of this, differential entropy ends up being a univariate function of *θθ*, for a specific set of parameters.

For the ranges of values typically found in the MMS, we see that Differential Entropy has almost a positive linear relationship with the natural logarithm of the scale, which is equivalent to the Noise-to-Signal Ratio of the gamma process. For greater values, that are not usually encountered in human data, the Differential entropy diverges. (Here the calculation of differential entropy for smaller values failed due to numerical instability). Figure 6A shows these results.

**Figure 6.**
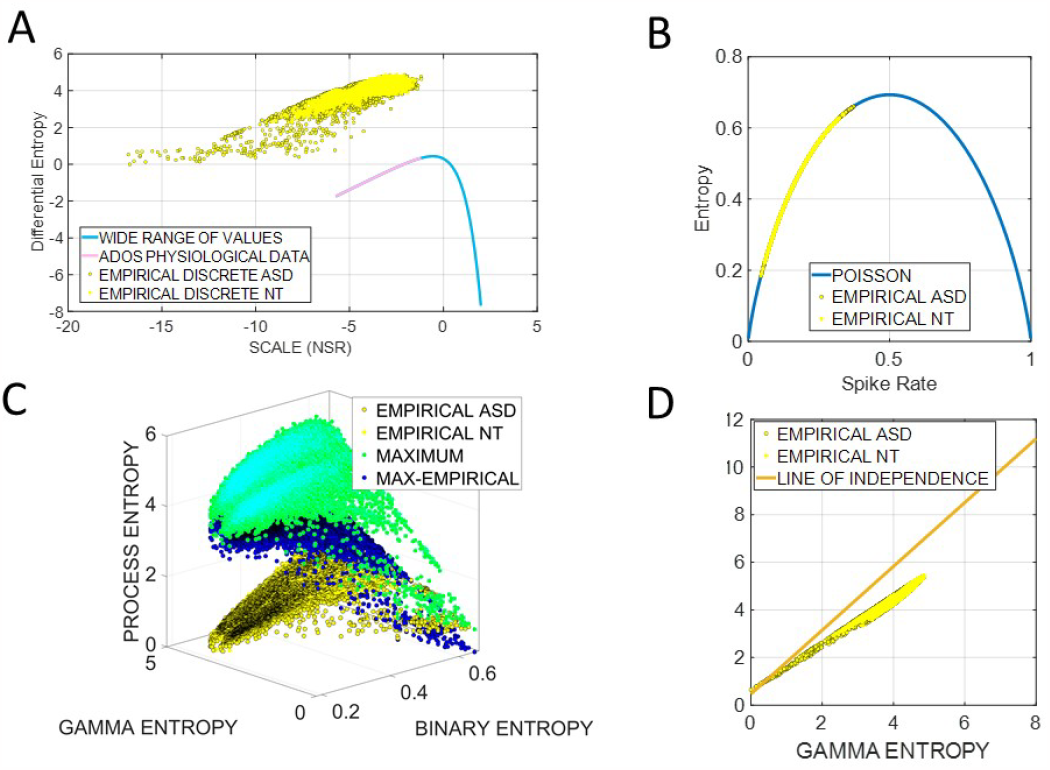
Theoretical considerations of relationships between NSR and Entropy metrics. (A) Near positive linear relationship between Differential Entropy and the natural logarithm of the Gamma scale, the NSR of the gamma process. (B) The empirically estimated entropy is the same as the equivalent Poisson process. Within the range of physiological data from the ADOS dyadic interaction, the entropy of the gamma component increases with the NSR, and the entropy of the binary component increases with the Spike Rate. (C) Sampling the empirical ranges across the entropy of the gamma component and the entropy of the binary component of the MMS yields the theoretical maximal entropy defining the upper bound (green). Their ratio (blue) indicates that the maximum entropy is greater than the empirical entropy by a quantity that is increasing as the binary entropy drops (*i.e*., as the spike rate decreases) and it increases as the gamma entropy increases. (D) The binary spike process has a relatively small dependence from the gamma process, suggesting that in human motion timing and amplitude (spatial) aspects of these motions are independent.

Because of Equation (4.10), we expect the empirical discrete entropy to also behave linearly with respect to the log-NSR, which turned out to be the case. For the range of physiological values, we found a slope of 0.4694 nats (p<0.01) for the differential entropy and a slope of 0.2606 nats (p<0.01) for the empirical entropy.

Moving on to the process *H*(*X*_*B*_), the theoretical entropy (if probabilities do not change within a time window, *i.e*., spike process is Bernoulli) is equal to [44]:

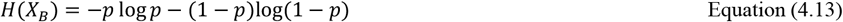

Where *p* is the probability of a spike occurring. We approximate it in Equation 4.14 as:

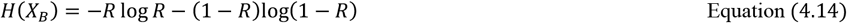

Where R is the spike rate measured as “number of spikes” per “number of samples”.

As we see, the empirically estimated entropy is the same as the equivalent Poisson process. As a conclusion, for the range of physiological data from the ADOS dyadic interaction, the entropy of the gamma component is increasing with the NSR, and the entropy of the binary component is increasing with the Spike Rate.

### 4.2 The Separability of Peak Activity from Standardized Angular Speed

If we plot the entropy of the process *H*(*X*_*G*_*X*_*B*_) and the maximum theoretical entropy defining the upper bound, *H*(*X*_*G*_) + *H*(*X*_*B*_), we see that the maximum entropy is greater than the empirical entropy by a quantity that is increasing as the binary entropy drops, or equivalently (by the previous finding) as the spike rate decreases. It increases as the gamma entropy increases. Fitting a surface function, we find that:

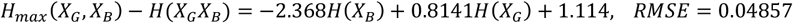

Then, we get an approximate relation:

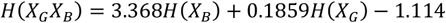

Recall that the following inequality holds:

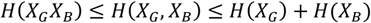

Which yields:

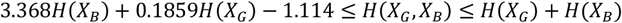

Or:

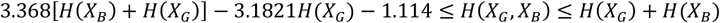

Finally:

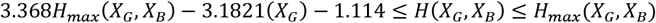

If *X*_*G*_ and *X*_*B*_ were to be independent, we can see that:

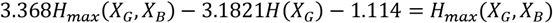

Or:

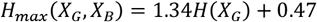

This independence criterion, is obviously data dependent. In the general case where we can fit a surface of the form:

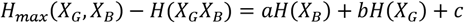

The condition for independence is:

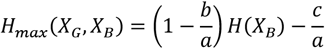

If the actual data can be well fit by a linear model with slope *A* and we ignore small differences between the intercepts of the two linear models, an easy way to quantify departure from independence is by computing the angle *θθ* between the two lines:

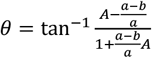

Then, we define the degree of departure from independence as the ratio between the *θ* and 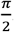. If the data cannot be well fit by a linear model, it’s best to perform a standard goodness-of-fit test and/or measure the mean error between the model and the data. If *θ* > 0, the rate of maximum joint entropy increase over the gamma entropy is bigger than in the case of independence, if *θ* < 0 it’s smaller.

From our data, we see that this condition holds pretty well, with 9.93 % departure from independence for the ASD group and 11.55 % for the NT group. This implies that:

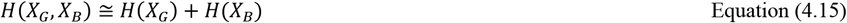

We just showed that the binary spike process is not very dependent from the gamma process. This supports the independence of time and space in human motion, previously proposed for intentional, goal-directed movements at a behavioral level of kinematic analyses [16; 45; 46; 47; 48] under a geometric modeling approach to address the brain control and coordination of the bodily degrees of freedom problem [49; 50].

### 4.3 Controllability of an Agent in a Dyadic Social Interaction is Inversely Proportional to Autonomy: Leveraging Sociomotor Agency to Protect the Agent

In the methods section we defined what Transfer Entropy *T*_*Y*→*X*_(*N, l*) between two processes X and Y is:

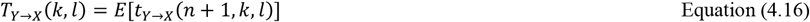

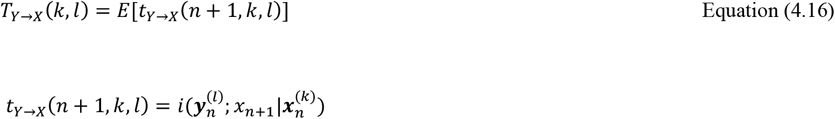

Equivalently, TE can be seen as the difference between the conditional entropy rate (which is equal to entropy rate for stationary processes) *h*_*X*_ of process X and the generalized entropy rate *h*_*X,Y*_ of X conditioning on the source Y [51]:

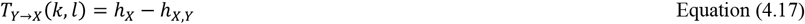

With:

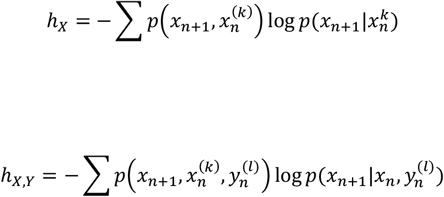

The generalized entropy rate measures the uncertainty in predicting the future values of X, given its history and the past values of Y. Transfer Entropy is the reduction in uncertainty of predicting the future of X when we consider the process Y. If we call *h*_*Y,X*_ uncertainty, then *h*_*Y*_ is what we already defined as autonomy and *T*_*X*→*Y*_ is the transfer entropy.

We chose embedded history of length 20 for TE and for the entropy rate of our processes we used a template (embedding) length equal to the average distance between two spikes, to ensure that in the reconstructed space, the coordinates of a point in time include both zeros (“quiet moments”) and spikes and that the system doesn’t bounce back and forth from a single coordinate of zeros components. The embedding delay was chosen using Average Mutual Information.

If we plot the Child or Clinician Autonomy with respect to the log(NSR) and the Spike Rate, we see in Figure 7 that the relationship between entropy rate, noise and spike rate is rather complex. It also differs between NT and ASD, more data are needed to get a clear picture but we can definitely see that there is a small positive trend with respect to noise and spike rate. Nonetheless, this shows that the processes cannot be treated as *i.i.d*.

**Figure 7.**
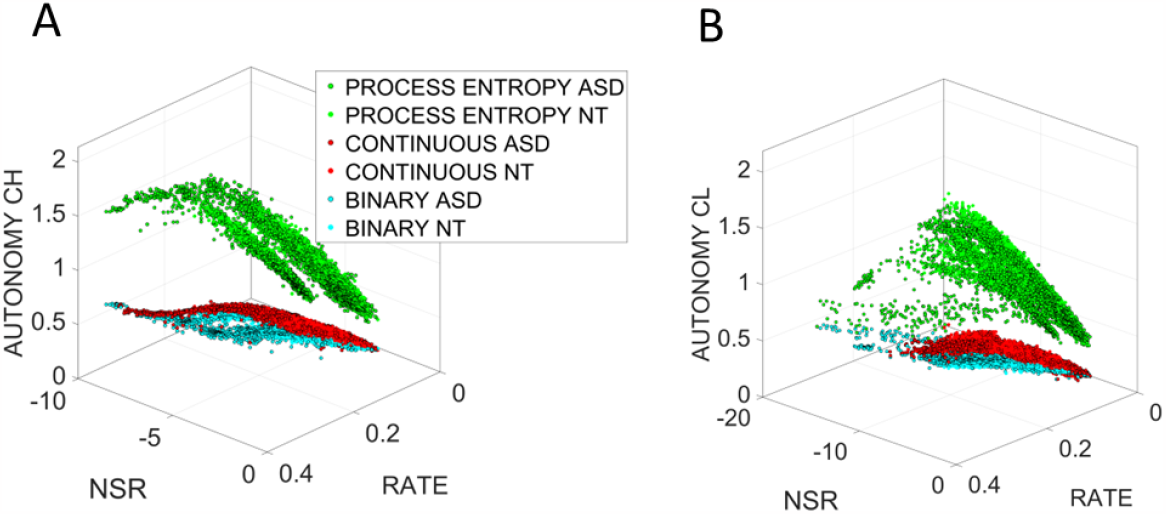
Non i.i.d. process revealed by the relationship between autonomy, NSR and spike rate for clinician (A) and child (B) for the gamma and binary components of the MMS, relative to the process entropy.

Now that we have established the speed/peak activity independence and the positive correlation between entropy rate and NSR or Spike Rate, we are ready to study how TE behaves in the shared space of the child-clinician dyad.

We find that *TE*_*CL*→*CH*_ decreases when the child exhibits high autonomy and increases when the clinician has higher autonomy and vice versa for *TE*_*CL*→*CH*_. In fact, this relationship is well characterized by linear relationships between transfer entropy and the entropy rates (autonomies), as the fitted linear surfaces indicate in Figure 8.

**Figure 8.**
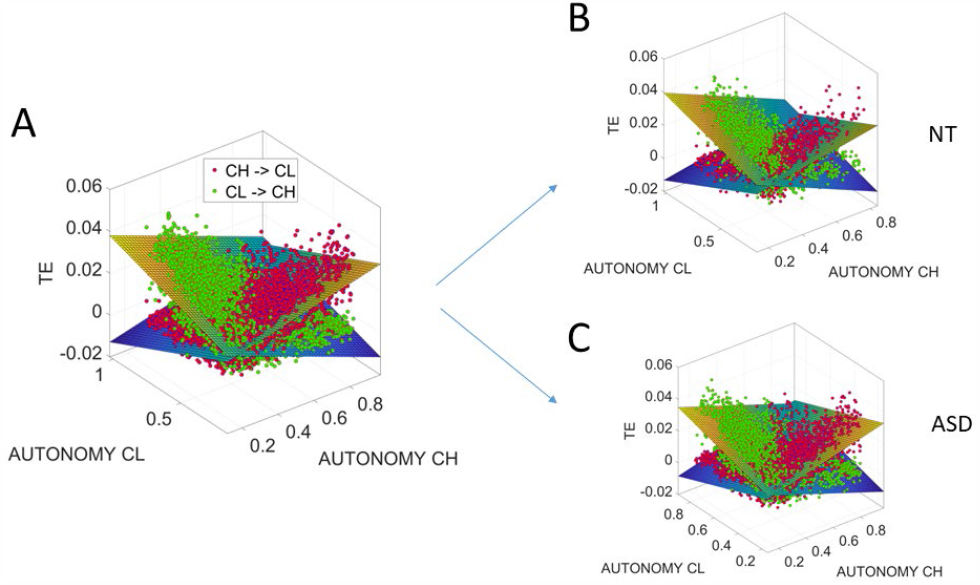
Linear relationships between transfer entropy and the entropy rates (autonomies) for child and clinician differentiating between NT and ASD participants.

In this sense, we can safely conclude that by manipulating standardized human biorhythmic time series either by increasing the NSR or by increasing peak activity, we can increase autonomy and reduce the controllability of human agents by other human or by artificial agents, including those potentially created by AI.

### 4.4 Validation of the Digitization of the ADOS: Automated, Streamlined and Scalable Screener of Socio-Motor Agency

To make our basic scientific results actionable, we need to validate our digital data with the clinical criteria, a paradigm that we have coined *clinically interpretable digital biomarkers*. In this model, the objective digital indexes that we used to define socio-motor agency as the autonomy-to-control ratio, are examined in relation to the ADOS clinical scores that a trained human rated during the session. We employ a machine learning technique, Support Vector Machine (SVM) to classify the digital data as a function of the clinical score. Then we apply tools from signal detection theory, specifically the receiving operating characteristic curve, ROC, to assess the validity of our classifier.

Each of 26 participants with the full ADOS session (digital and clinical) produces on average between 50 – 60 minutes of time series digital data from biosensors registering motion at 128Hz. We used the left-hand wrist sensor in these analyses, as we showed that it is highly correlated with the right wrist, yet more variable, thus expanding our sampling space. Upon exploration of several time windows to segment the data, sweeping across the time series and tasks, while maximizing statistical power in each locally stationary segment, we arrived at 7.8 second windows as optimal.

The data were validated using the Leave-one-person-out cross-validation (LOOCV) method. As features for our classifier, we used autonomy (entropy rate), NSR and the embedding delay of the data, which is the time scale at which deterministic properties arise and characterize the dynamical behavior of motion (for more information, see Methods). Two classifiers were used, one trained on female subjects and the second one trained exclusively on male participants. When trying to digitally diagnose autism in one participant, we trained our classifier on the data from the remaining male or female participants and then tested how accurately the trained model predicts the participant class (NT *vs*. ASD). This method avoids overfitting and trains models that can diagnose autism in novel participants, thus automating the screening process. Digitizing the ADOS in this way makes the diagnosis of autism more inclusive of females, historically underdiagnosed by a test that we objectively showed has biases towards males across all tasks [10]. A larger sample size and a longitudinal study are required to validate our model at scale. Yet, as shown in Figure 9A, there is no confusion of our biometrics about the clinician ADOS scores, which classify ASD males with 100% accuracy and performs remarkably well for ASD *vs*. NT females. Indeed, Figure 9B confirms the validity of these biometrics for clinical use with an area under the ROC curve of 95.76%.

**Figure 9.**
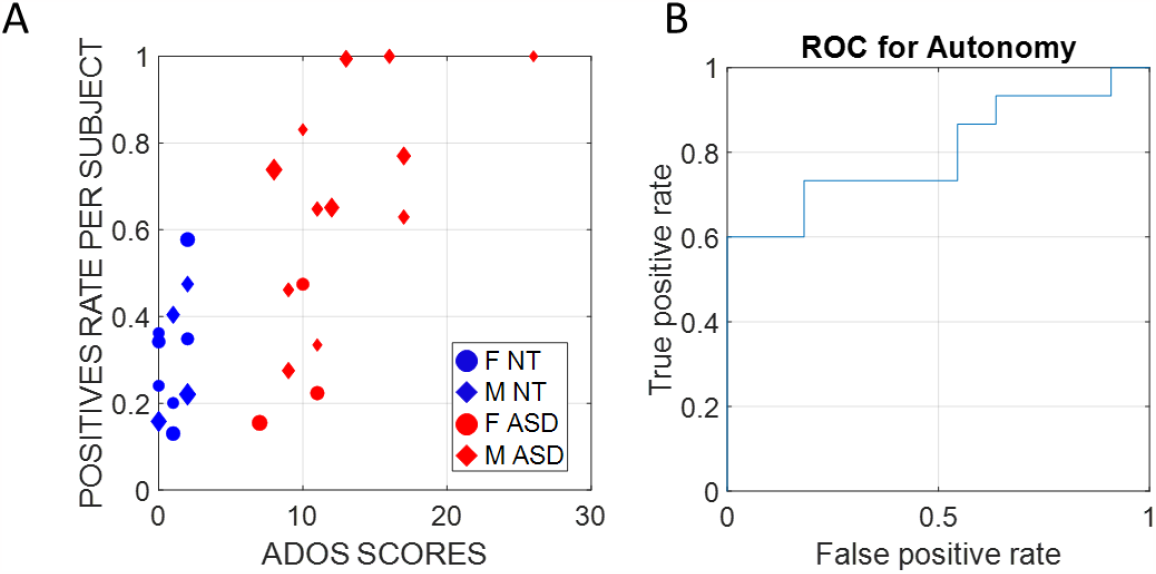
(A) Support Vector Machines (SVM) classifiers were trained on all subjects except one and tested on the remaining subjects of the same sex (Leave-one-person-out cross-validation (LOOCV)). Therefore, each of the 26 subjects was digitally diagnosed with a classifier trained on a different dataset, which ensured zero overfitting and bias. Training and testing features were the entropy rate, the signal-to-noise ratio, and the embedding delay (the time scale at which a dynamical system behaves in the most deterministic way) calculated on normalized speed samples of ∼7.8 secs duration windows. Here, we report the percentage of time windows per subject that gave a positive diagnostic label and plot them versus the ADOS scores, as determined by the clinicians. (B) We use the positive rate scores as a metric used to diagnose ASD and report the Receiver Operating Characteristic curve (ROC curve), which shows the true positive and false positive rates of the digital diagnostic tool we developed for different thresholds. The Area Under the Curve (AUC) is 0.9576, which indicates great performance.

## 5 Discussion

In this work, we use the ADOS test as a backdrop to study social interactions between children and adult clinicians with the purpose of defining new ways to automate and speed up the autism screening process, while leveraging the clinical validity of this test. To that end, we explored anew the concept of socio-motor agency by defining a ratio of two indexes of autonomy and control. Autonomy was defined as the non-parametric entropy rate spanning from totally random to totally deterministic behavior of standardized micro-movements spike trains. These were derived from nuanced fluctuations in motion data that contains goal-directed segments of behavior interspersed with spontaneously occurring, more ambiguous, transient segments that are known to interconnect the goal-directed ones [16; 52]. Control was defined in terms of the NSR, empirically estimated from such spike trains as well, such that high regimes of NSR correspond to the memoryless regimes denoting high uncertainty (poor predictability and randomness) in the motor code. We took a step further to examine the parameterization of the MMS as a binary-spike and a Gamma process and demonstrated the independence between them.

We reasoned that these binarized sequences of spikes bear a motor code whereby the observer may or may not be able to predict and therefore control the observed agent. At high randomness, the observed agent affords more autonomy than at deterministic ranges. At deterministic ranges, with high regularity, the observer can predict and control the actions of the observed agent. At higher NSR, the agent has lower self-control. This is so because the kinesthetic reafferent feedback from the motions is noisy and with such poor signal quality it is difficult to predict a desired outcome and plan the action consequences to compensate for sensory transduction, transmission, and motor integration delays inherent in the person’s system. As predicting his/her/their motor actions consequences can then be compromised by noise in the motor code, the child is more controllable by the clinician. The observer clinician can exert higher control over the observed agent. In this sense, the child’s socio-motor agency may also be compromised. This is the case whether the child / adult is autistic.

Underlying both indexes and the ratio of autonomy to control are then discrete pockets of information making up a continuous stream of dyadic motor code, contributed by both social agents. Thus, we can infer the existence of an underlying shared alphabet in the motor code that manifests during dyadic social interactions of the type studied here. Agents with discrete motor signatures that appear more random are thus harder to control and behave more autonomously and independently than agents with systematically predictable motions sharing their codes.

Besides describing new biometrics of shared socio-motor agency in dyadic social interactions, our analyses showed ways to streamline the ADOS test, thus making it less taxing on the child and the clinician. A handful of tasks affording more socio-motor agency to the child can indeed uncover the social readiness potential of the child rather than biasing the diagnosis by the clinician towards a deficit model. Along those lines, using these newly defined indexes of dyadic autonomy and control, we demonstrated fundamental differences across the tasks for males and females, thus confirming that despite previously quantified differences in motor control separating males and females at the voluntary [4] and involuntary [12; 13] levels, the ADOS remains biased towards males. These digital indexes of shared socio-motor agency, nevertheless, used within the context of an unbiased ML classifier, could detect the differences between males and females for both the NT and ASD randomly chosen participants. This digitized automated version of the test resembles the type of scenario that a clinician faces at the clinic, any given day. Namely, a random arrival of a case that the clinician may see for the first time. In that sense, the leave-one-person-out classifier provides robust digital screening of autism and may be a way to scale our pilot study to encompass larger numbers of NT, ASD participants across ages, sexes, and do so longitudinally as well.

Future longitudinal studies of autism with an eye for the evolution of the neuromotor code and its impact on social perception and cognition, will require the type of normalization that we introduced earlier with the MMS [3] and further used here, namely, scaling out allometric effects due to anatomical differences across participants (see also [17; 18; 53; 54]). This step is crucial in any study that involves biorhythmic motions whereby kinematic analyses will be impacted by such anatomical differences. This is so because kinematic parameters such as speed, acceleration, distance, etc. are impacted by the limb sizes and masses in ways that confound results and interpretation of such studies [55]. It will be particularly important to consider these caveats present in all current studies that do not account for allometric differences during the very early neurodevelopment when the rate of bodily growth is highly non-linear and accelerated [54]. These rates of changes in anatomical growth produce different ranges of values in such kinematic parameters and impact the empirical distributions of the values associated with natural behaviors such as those examined here.

### 5.1 Implications of Socio-Motor Agency Metrics for AI and Privacy Protection

The theoretical considerations at the intersection of stochastic analyses and information theoretic approaches with non-linear dynamics offers the MMS and analyses as a viable way to obtain the personalized signatures of autonomy and control and tweak the NSR to mask the spike trains derived from the person’s physiological biorhythmic activity. This ability to separate the binary spike rate code from the gamma process denoting levels of randomness *vs*. predictability, offers the possibility of creating a device that alerts the persons involved in the dyadic exchange to balance their autonomy and control, to attain socio-motor agency. By enhancing autonomy and avoiding excessive external control by the other agent, be that agent another human or an AI-driven one, the person can be protected from excess control. This approach will be critical to revamp autism therapies with an emphasis to respect the child’s autonomy and support the bottom-up development of autonomous motor control. The maturation of bottom-up autonomous motor control (building blocks of autonomy) is a necessary pre-requisite for the further neurodevelopment of top-down control. Without considering and balancing the orderly maturation rates of these two building blocks of socio-motor behavior, therapies in autism will cause trauma to the nervous system.

We propose that this methodology can also be used to protect our privacy more generally from surveillance systems, as ultimately these systems rely on biometric data, which we can now, using the present personalized methods, manipulate to hide our fingerprint-like signatures from an external agent trying to control us. This solution to the controllability issue can then be extended from individuals to dyads, from dyads to social groups and from social groups to society. In this sense, socio-motor agency can serve as a foundation for societal agency, now quantifiable using the methods that we offer in this work.

## 6 Conclusions

In summary, we found that variability in the dyadic index of autonomy is more pronounced in ASD than in NTs, across a broad range of ages from 4-15 years old. Furthermore, we found that the dyadic NSR, indicative of socio-motor control, increases with age. This result is consistent with prior work on individuals across ages and sex [3; 4]. In contrast, both ASD and NT showed increases of the autonomy index with age, an indicator that regardless of the human condition, whether developing along a neurotypical trajectory, or along the trajectory of autism spectrum disorders, respecting the child’s autonomy will be necessarily our best ally when designing future treatments that unveil the child social readiness potential. We would not have known this had we treated the ADOS as the criterion test that it is (*i.e*., based off children with neurodevelopmental issues only), rather than treating it as a normative test (*i.e*., including NT controls as well, to define normative ranges and quantify similarities and departures from it.)

We have uncovered new indexes of shared, dyadic autonomy and control, objectively defined socio-motor agency and provided new means to automate its digital screening with already routinely used clinical tools. This works offers novel ways to scale our clinical science and make it actionable, diverse, and inclusive at more than one level.

## Data Availability

All data produced in the present study are available upon reasonable request to the authors
All data produced are available online upon request. Link in the Data Availability section of the manuscript.

https://zenodo.org/records/10032169

## 8 Author Contributions

TB and EBT contributed to conception and design of the analyses. TB analysed the data while EBT designed the clinical study. RR organized and curated the database. TB wrote methods and derivations. EBT wrote report. TB and EBR wrote and edited full manuscript. All authors contributed to manuscript revision, read, and approved the submitted version.

## 10 Acknowledgments and Disclaimer

We thank the children and families who kindly participated in the study. **This study was approved by the Rutgers University IRB and signed consent was obtained from the legal guardian/parent of the child**.

## 12 Data Availability Statement

The datasets [GENERATED/ANALYZED] for this study can be found in the repository https://zenodo.org/records/10032169. Please see the “Availability of data” section of [Materials and data policies in the Author guidelines](https://www.frontiersin.org/guidelines/policies-and-publication-ethics#materials-and-data-policies) for more details.

